# Validity of the NINDS traumatic encephalopathy syndrome criteria for predicting chronic traumatic encephalopathy

**DOI:** 10.64898/2026.05.24.26353505

**Authors:** Jesse Mez, Bobak Abdolmohammadi, Shruti Durape, Brigid Dwyer, Michael L. Alosco, Belinda Yew, Alexandra Pritchett, Natalia Bernal Fernández, Amelia Hicks, Madeline Uretsky, Megan Ryder, Farwa Faheem, Sophia Nosek, Brett Martin, Joseph Palmisano, Christopher Nowinski, Yorghos Tripodis, Kristen Dams O’Connor, Lee E. Goldstein, Douglas I. Katz, Robert C. Cantu, Neil W. Kowall, Robert A. Stern, Victor E. Alvarez, Bertrand R. Huber, John F. Crary, Thor D. Stein, Ann C. McKee, Daniel H. Daneshvar

**Affiliations:** Boston University Alzheimer’s Disease Research Center and CTE Center, Boston University Chobanian and Avedisian School of Medicine, Boston, MA, USA; Department of Neurology, Boston University Chobanian and Avedisian School of Medicine, Boston, MA, USA; Department of Neurology, Boston Medical Center, Boston, MA, USA; Framingham Heart Study, Boston University Chobanian & Avedisian School of Medicine, Boston, MA, USA; Braintree Rehabilitation Hospital, Braintree, MA, USA; Department of Anatomy & Neurobiology, Boston University Chobanian & Avedisian School of Medicine, 72 East Concord St., Boston, MA 02118, USA; Department of Rehabilitation and Human Performance, Brain Injury Research Center, Icahn School of Medicine at Mount Sinai, New York, NY USA; Department of Neurology, Icahn School of Medicine at Mount Sinai, New York, NY USA; Department of Neuroscience, Icahn School of Medicine at Mount Sinai, New York, NY USA; Department of Psychological Sciences, University of Connecticut, Storrs, CT USA; Department of Psychology, University of British Columbia, Vancouver, BC, Canada; Biostatistics and Epidemiology Data Analytics Center, Boston University School of Public Health, Boston, MA, USA; Concussion and CTE Foundation, Boston, MA, USA; Department of Biostatistics, Boston University School of Public Health, Boston, MA, USA; Departments of Biomedical, Electrical & Computer Engineering, Boston University College of Engineering, Boston, MA, USA; Department of Psychiatry, Boston University Chobanian and Avedisian School of Medicine, Boston, MA, USA; Department of Radiology, Boston University Chobanian and Avedisian School of Medicine, Boston, MA, USA; Department of Neurosurgery, Emerson Hospital, Concord, MA, USA; Department of Neurosurgery, Boston University Chobanian and Avedisian School of Medicine, Boston, MA, USA; VA Boston Healthcare System, U.S. Department of Veteran Affairs, Boston, MA, USA; VA Bedford Healthcare System, U.S. Department of Veteran Affairs, Bedford, MA, USA; Uniformed Services University of the Health Sciences, Bethesda, MD, USA; Departments of Pathology, Molecular & Cell-Based Medicine, Neuroscience, and Artificial Intelligence & Human Health, and the Neuropathology Brain Bank & Research Core, Icahn School of Medicine at Mount Sinai, New York, NY, USA; Department of Pathology and Laboratory Medicine, Boston University Chobanian and Avedisian School of Medicine, Boston, MA, USA; Department of Rehabilitation Medicine, Harvard Medical School, Boston, MA, USA; Department of Physical Medicine and Rehabilitation, Massachusetts General Hospital, Boston, MA, USA; Department of Physical Medicine and Rehabilitation, Spaulding Rehabilitation Hospital, Boston, MA, USA

## Abstract

**Importance:** Accurate prediction of chronic traumatic encephalopathy (CTE) remains challenging in life.

**Objective:** To assess the reliability and validity of the NINDS traumatic encephalopathy syndrome (TES) criteria to predict CTE pathology in life.

**Design:** Clinicopathological Diagnostic/Prognostic Study

**Setting:** Six brain banks with varied recruitment criteria

**Participants:** Brain donors were selected across 6 brain banks (15+ donors each), 5 age groups spanning ages 20 to 80+ (25+ donors each) and 9 repetitive head impact (RHI)/traumatic brain injury (TBI) groups (15+ donors each): (1) college or professional American football; (2) less than college football; (3) college or professional contact sports, non-football; (4) less than college contact sports, non-football; (5) military combat, no contact sports; (6) military combat and contact sports; (7) concussion with loss of consciousness, no RHI; (8) moderate to severe TBI, no RHI; (9) no RHI/TBI.

**Exposures:** Blinded to neuropathological information, clinicians reviewed prospective study and medical records and conducted informant interviews, and an expert panel adjudicated TES diagnoses, including provisional levels of certainty for CTE pathology (suggestive/possible/probable). TES diagnoses were *a priori* dichotomized: TES with possible/probable CTE (CTE_pos/prob_) vs. no TES/TES with suggestive CTE (CTE_sug_).

**Main Outcomes and Measures:** Blinded to clinical information, neuropathologists applied NINDS/NIBIB CTE neuropathological criteria and staging (I-IV). CTE diagnoses were *a priori* dichotomized: stages II-IV vs. no CTE/stage I.

**Results:** Among 193 brain donors [men:153 (79.3%), mean age:66.4 (SD:22.0)], 57 (29.5%) donors met clinical criteria for CTE_pos/prob_ and 42 (21.8%) donors met neuropathological criteria for CTE stages II-IV. There was high agreement between panelists for CTE_pos/prob_ vs. no TES/CTE_sug_ (ICC:0.95, 95%CI:0.88-0.97). CTE_pos/prob_ sensitivity, specificity, positive likelihood ratio (LR) and negative LR for CTE stages II-IV were: 0.77 (95%CI:0.64-0.89), 0.84 (95%CI:0.78-0.90), 4.8 (95%CI:3.02-7.61), 0.28 (95%CI:0.15-0.50); age≥50:0.90 (95%CI:0.80-1), 0.90 (95%CI:0.85-0.96), 9.2 (95%CI:4.9-17.27), 0.11 (95%CI:0.04-0.33). All younger false positives (age<50; n=13) had a mental health, substance use and/or pain disorder. All older false positives (age≥50; n=11) had non-CTE neurodegenerative and vascular pathologies. Among 10 false negatives, 8 had stage II CTE.

**Conclusions and Relevance:** The NINDS TES criteria demonstrated good reliability, sensitivity and specificity, and provided moderate to large evidence to both rule out and rule in CTE pathology, particularly above age 50.

**Key Points:** *Question:* What is the validity of the NINDS consensus diagnostic criteria for traumatic encephalopathy syndrome (TES) for predicting chronic traumatic encephalopathy (CTE) neuropathology?

*Findings:* In this clinicopathological diagnostic/prognostic study that included brain donors from varied brain banks, head impact exposures and ages, TES criteria sensitivity, specificity, positive likelihood ratio (LR) and negative LR were 0.77, 0.84, 4.8 and 0.28 with improved performance above age≥50 (0.90, 0.90, 9.2, 0.11).

*Meaning:* The NINDS TES criteria were sensitive and specific for CTE neuropathology across varied head impact exposures, particularly above age 50.

## Introduction

Chronic traumatic encephalopathy (CTE) is a neurodegenerative tauopathy most commonly found in individuals with exposure to repetitive head impacts (RHI),^1–7^ including former contact and collision sport athletes and military veterans.^2–4,6,8,9^ CTE can only be definitively diagnosed by post-mortem neuropathologic examination. A consensus panel of neuropathologists organized by the National Institute of Neurological Disorders and Stroke and the National Institute of Biomedical Imaging and Bioengineering (NINDS-NIBIB) defined and refined criteria for chronic traumatic encephalopathy (CTE) diagnosis.^10,11^ The panel concluded that CTE is a unique tauopathy reliably distinguished from other neurodegenerative diseases.

The clinical syndrome associated with CTE is less clearly defined. Clinical features of CTE have been described in larger restropective and small prospective reports of brain donors with neuropathologically confirmed CTE.^5,12–15^ Among 111 former American football players diagnosed with CTE, cognitive, behavioral, and mood symptoms were commonly reported by informants, as was dementia and a progressive clinical course.^5^ Several prospective and retrospective studies have described memory and executive function decline in the presence of CTE pathology, often together with other neurodegenerative pathologies.^5,13–15^ Among 364 brain donors with autopsy-confirmed CTE, regional phosphorylated tau (ptau) burden, particularly in the frontal cortex, was clearly associated with severity of informant-reported cognitive and functional symptoms, with far weaker association with neurobehavioral dysregulation.^16^ Among 615 brain donors with (n=366) and without (n=248) autopsy-confirmed CTE without other neurodegenerative pathologies, dementia was 4.5 times more likely in donors with stage IV CTE compared to those without CTE.^17^

Multiple iterations of diagnostic criteria for the clinical syndrome associated with CTE pathology have been proposed.^18–21^ In 2014, research diagnostic criteria for traumatic encephalopathy syndrome (TES) were proposed to “describe the clinical presentation of CTE as well as other possible long-term consequences of repetitive head impacts”.^22^ Retrospectively applied to 336 RHI-exposed brain donors to the Understanding Neurological Injury and Traumatic Encephalopathy (UNITE) Brain Bank, the criteria demonstrated high sensitivity (0.97), but low specificity (0.21) for predicting CTE pathology.^23^ Other studies have similarly suggested that the 2014 criteria were not sufficiently specific.^24,25^ To improve upon the performance of the initial TES diagnostic criteria, 20 clinician-scientists participated in a four-round modified Delphi consensus process, resulting in the 2021 National Institute of Neurological Disorders and Stroke (NINDS) diagnostic criteria for TES.^26^ The panel came to consensus on four primary diagnostic criteria for TES, as well as several supportive features. These primary and supportive criteria informed provisional levels of certainty for CTE pathology (suggestive, possible and probable), based on a stepwise assessment of source and level of RHI exposure, specific clinical features, and a set of supportive features.^26^

Subsequently, an NINDS U54 grant was funded to leverage multiple brain banks, each with different enrollment criteria and demographic representation, to investigate contributions of traumatic brain injury (TBI) and RHI to Alzheimer’s disease and related dementias (AD/ADRD) neuropathology, including CTE, and resulting clinical outcomes. One of the grant aims and the goal of the current study was to evaluate the performance of the NINDS TES criteria against gold-standard CTE neuropathology. To maximize generalizability, a panel of interdisciplinary clinical experts blinded to pathology applied the NINDS TES criteria to a diverse group of brain donors selected with deliberate representation across six brain banks, five age groups spanning ages 20 to 80+, and nine head impact exposure groups spanning different forms of RHI and TBI. All cases underwent a full neuropathological evaluation, including for CTE diagnosis and stage, by an expert panel of neuropathologists blinded to clinical presentation. We hypothesized that the NINDS criteria would demonstrate improved specificity, while maintaining similar sensitivity compared with the initial 2014 criteria.

## Methods

### Study recruitment and selection

We selected brain donors based on recency of donation, representation across head impact exposure groups, age groups and brain banks and availability of neuropsychological testing. Had selection been based purely on recency of donation, these additional variables, which we deemed important for generalizability, would not have been sufficiently distributed across donors. A Standards for the Reporting of Diagnostic Accuracy Studies (STARD) flow diagram of included and excluded donors is shown in **eFigure 1**.

We required a minimum of 15 donors from each of the 6 brain banks:

1. UNITE^27^
2. Framingham Heart Study (FHS)^28^
3. Boston University Alzheimer’s Disease Research Center (BU ADRC)^29^
4. Mount Sinai Alzheimer’s Disease Research Center (Mount Sinai ADRC)^30^
5. Veterans Affairs Amyotrophic Lateral Sclerosis Biorepository (VA ALS)^31^
6. Late Effects of Traumatic Brain Injury (LETBI)^32^

Details of each brain bank are described in the **eMethods** in Supplement 1. We required a minimum of 25 donors from each of the 5 age groups:

1. 20-34 years old at death
2. 35-49 years old at death
3. 50-64 years old at death
4. 65-79 years old at death
5. 80+ years old at death

We required a minimum of 15 donors from each of the 9 RHI/TBI exposure groups:

1. No RHI or TBI history
2. Mild TBI with loss of consciousness, no other RHI history^33^
3. Moderate-severe TBI, no other RHI history^34^
4. Military service with combat, and no contact sport play
5. Military service with combat, and contact sport play
6. American football play, less than college level
7. American football play, at least college level
8. Contact sport play (other than American football), less than college level
9. Contact sport play (other than American football), at least college level

### Neuropathological assessment

Across all brain banks, for each donor, a designated neuropathologist (ACM, TDS, BRH, JC) performed a comprehensive work-up blinded to the donor’s brain bank membership (within, but not across institutions) and RHI and clinical history. Neuropathological processing and examination followed previously detailed methods.^35,36^ Paraffin-embedded tissue sections were stained for Luxol fast blue, hematoxylin and eosin, Bielschowsky silver, ptau (AT8), a-synuclein, amyloid-β, and phosphorylated transactive response DNA binding protein 43 kDa (pTDP-43).^36^ CTE was diagnosed using NINDS/NIBIB neuropathological criteria.^11^ Donors with CTE pathology were also classified into low and high severity and 4 distinct stages (I-IV) using previously validated McKee staging scheme.^1,7,11^ Established criteria were used for diagnosing Alzheimer’s disease (AD),^37^ Lewy body disease,^38^ frontotemporal lobar degeneration (FTLD-TDP43, FTLD-tau, including progressive supranuclear palsy (PSP), corticobasal degeneration (CBD), Pick’s disease, and argyrophilic grain disease (AGD)),^39^ Limbic-predominant Age-related TDP-43 Encephalopathy (LATE)^40^ and motor neuron disease.^41^ Neuropathological examination also assessed various cerebrovascular pathologies, including arteriolosclerosis, atherosclerosis, white matter rarefaction and cerebral amyloid angiopathy, scored on a 0-3 scale, with 3 representing severe burden. Presence of gross and microscopic infarcts was also assessed.

### Collection of demographics, RHI, and clinical data

All clinicians and research assistants involved with clinical data collection and review were blinded from neuropathological results. A subset of donors were part of prospective research studies (BU ADRC, FHS, Mount Sinai ADRC, LETBI, VA ALS), and information from these donors during life was assembled and reviewed. For cases that did not already have medical records on hand, formal medical record requests were made following a previously described protocol.^27^ In addition to review of prospective data and medical records, data was also collected from informants. Questionnaires administered to informants collected information on donors’ demographics (race, sex, education), athletic history for each contact sport played (level of play, position(s) of play, age of first exposure, and duration of play), military history (branch, combat exposure, duration of service), cause of death and medical, TBI, neurological, psychiatric, substance use, social and family histories.^5,42–44^ This data ascertainment was standardized across all brain banks using NIH Common Data Elements to the extent possible given differences in study design and study-specific requirements about informant burden. **eTable 1** provides details about questionnaires administered for each brain bank. For each donor, phone interviews with informants were conducted by a lead clinician (JM, BD, MA, BY, AP, NBF, AH, DHD) with expertise in neurodegenerative diseases. Utilizing an unstructured interview format, clinicians obtained a detailed timeline of cognitive, mood, behavioral and motor symptoms, as well as sleep, headache and pain.

After completion of prospective study record review, medical record review, and informant interviews, the lead clinician qualitatively summarized participants’ clinical timelines into a narrative summary suitable for presentation at a multidisciplinary clinicopathological consensus conference. This summary included the age and cause of death, the clinician’s subjective evaluation of the informant’s reliability using a three-point scale (2: very reliable, i.e., could answer most questions knowledgeably, and consistent with prospective data and medical records; 1: somewhat reliable, i.e., could answer some questions knowledgeably, and consistent with prospective data and medical records; 0: unreliable: could answer few questions knowledgeably, and consistent with prospective data and medical records), history of RHI, clinical trajectory, prior medical, educational, and occupational backgrounds, living arrangements prior to death, substance-use history, family history and relevant medical record information. Additional clinical data, such as results of neuropsychological assessments, neuroimaging (with expert interpretation when primary images were accessible), cerebrospinal fluid biomarkers of AD, antemortem diagnoses and medication histories were also included.

### Clinicopathologic consensus conferences

Clinicopathologic consensus conference (CPC) methodology followed previously published guidance.^45^ CPCs began in April 2021 and continued through May 2024. A panel of clinicians (neuropsychologists, neurologists, psychiatrists, a physiatrist, and a neurosurgeon; JM, BD, MA, BY, AP, NBF, AH, KDOC, LG, DIK, RCC, NWK, RAS, DHD) with expertise in neurodegenerative disease met twice monthly. Each CPC required at least three panel members with at least 1 neurologist and 1 neuropsychologist (mean: 8.4 panel members). Both the lead clinician and participating clinician panelists were blinded to neuropathological findings until after voting was concluded. For each case under review, the lead clinician distributed and presented aloud to the panel the prepared summary (structure previously described). Following presentation, and prior to group discussion, panelists independently entered into an online form item-level responses including TES primary and supportive features used to determine whether the NINDS TES criteria were met^23,26^ and, if so, the provisional level of certainty for CTE: suggestive (CTE_sug_), possible (CTE_pos_), or probable (CTE_prob_)], as well as additional diagnoses contributing to the clinical presentation (**eMethods**, **eFigure 2**). Once forms were submitted, an electronic report that compiled the responses was generated identifying which components had clear group majority consensus and which required further discussion to resolve. Group discussion ensued, including soliciting additional details from the lead clinician, with the objective of achieving a majority consensus for each criterion. Whether a consensus TES determination was made, including the provisional level of certainty of CTE, and additional diagnoses, were determined based on post-discussion voting. Next, the neuropathologist assigned to each case (ACM, TDS, JC, BRH) disclosed the pathological diagnoses, presenting gross and microscopic neuropathological findings.

### Analyses

Prior to study onset, we chose the primary clinical diagnostic cut point to be TES with possible or probable level of CTE certainty (CTE_pos/prob_) vs. no TES or CTE_sug_. CTE_sug_ was not grouped with CTE_pos/prob_ to increase likelihood that the criteria would be sufficiently specific. However, sensitivity analyses included alternative groupings: 1) TES with all levels of CTE certainty (CTE_sug/pos/prob_) vs. no TES and 2) CTE_prob_ vs. no TES or CTE_sug/pos_. We chose for the primary CTE neuropathological cut point to be stages II-IV vs. no CTE or stage I. Stage I was not grouped with stages II-IV because early CTE tau pathology has not been consistently associated with clinical symptoms.^46^ Stage II was grouped with stages III-IV to assure that the positivity assumption was met with respect to young age.^47^ Young donors rarely have stages III and IV pathology and, if Stage II were not grouped with Stages III and IV, young donors would not be able to have the outcome of interest.^46^ Similarly, sensitivity analyses included alternative groupings: 1) all stages of CTE pathology vs. no CTE pathology and 2) CTE stages III-IV vs. no CTE pathology or CTE stages I-II.

#### Interrater reliability

Analyses to assess TES interrater reliability used diagnoses made by clinician panelists <u>prior</u> to discussion to reach a consensus. Interrater reliability was computed using intraclass correlations (ICC) estimated by generalized linear mixed effects models with a logit function. We used ICC in order to account for variability in voters across cases, under a missing at random assumption.^23,48^ 95% confidence intervals were generated with bootstrapping. Primary and sensitivity analyses used dichotomizations for TES diagnoses as described above.

#### Validity

Analyses to assess TES validity used diagnoses made by clinician panelists <u>after</u> discussion to reach a consensus. Sensitivity, specificity, positive likelihood ratio (LR+) and negative likelihood ratio (LR-) were calculated based on standard definitions using CTE neuropathology as the gold standard. Primary and sensitivity analyses used dichotomizations for TES diagnoses and CTE neuropathologic diagnoses as described above. Additional stratified sensitivity analyses were also performed to investigate if certain hypothesized factors affected TES performance: age at death at 50 years of age (chosen to include the recruitment age bin that includes most early-onset dementia, i.e., 50-65), presence of formal neuropsychological testing, presence of clinician panel diagnoses of DSMV substance use disorder or mental health disorder (defined as DSMV diagnoses of major depressive disorder, bipolar disorder, anxiety disorders, trauma and stressor-related disorders, schizoaffective disorder, schizophrenia, or other psychotic disorder), and presence of neuropathological diagnosis of Alzheimer’s disease (intermediate or high). To investigate if the clinical features improved performance beyond the RHI exposure cut point, additional sensitivity analyses were performed just using the cut-point for “substantial” RHI exposure alone (as defined by the NINDS TES criteria) without any clinical features from the criteria or using both the cut-point for substantial RHI exposure and the core feature of cognitive impairment, but without considering other clinical features.

Lastly, using a chi squared test, we investigated whether among donors with substantial RHI exposure, the accuracy of CTE_pos/prob_ for predicting CTE stages II-IV differed by whether AD pathology was present (defined by Reagan criteria for possible or probable likelihood of dementia due to AD), as it has well-established biomarkers and had associations with TES accuracy for the 2014 criteria.^23^

#### Item level data exploration

False positive and false negative cases from the validity analyses were explored and presented graphically. Frequencies of donors with CTE stages II-IV were presented in Venn diagrams with groupings by item level TES primary diagnostic features to investigate if certain features had outsized contributions towards an accurate determination.

Data were collected, stored, and managed using REDCap electronic data capture tools hosted at Boston University, CTSI 1UL1TR001430. Statistical analyses were conducted using SPSS Statistics version 29.0.1.0, and R 4.5.0.

## Results

Among 193 total donors [mean age at death: 66.3 years (standard deviation (SD): 22.0); female: 40 (20.7%)], 72 donors were diagnosed with TES (37.3%), including 15 with CTE_sug_ (7.8%), 28 with CTE_pos_ (14.5%), and 29 with CTE_prob_ (15%). Fifty-seven donors had CTE pathology (29.5%), including 14 with stage I (7.3%), 11 with stage II (5.7%), 16 with stage III (8.3%) and 16 with stage IV (8.3%). A total of 57 donors were diagnosed with CTE_pos/prob_ (29.5%) and 43 donors had CTE stages II-IV (22.3%), the dichotomizations used in the primary analyses described below. **Table 1** shows demographics, donor recruitment categories, item level components of the TES criteria (as voted on by the clinician panel), availability of biomarkers and neuropathologies stratified by TES diagnostic categories. There was an average of 8.4 panel voters (SD: 1.8) per case. The average informant reliability score was 1.83 out of 2 (SD: 0.37).

**Table 1.**
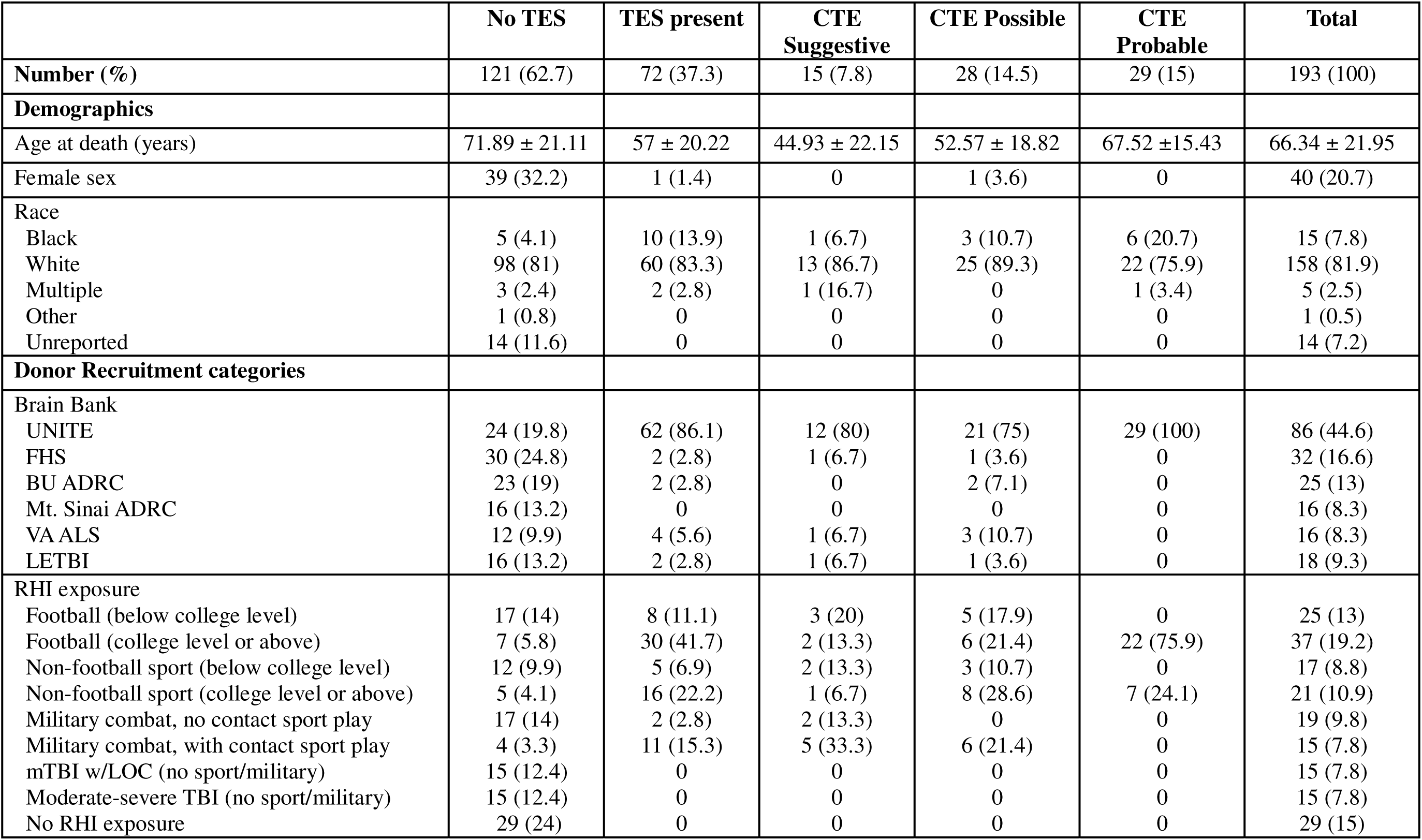
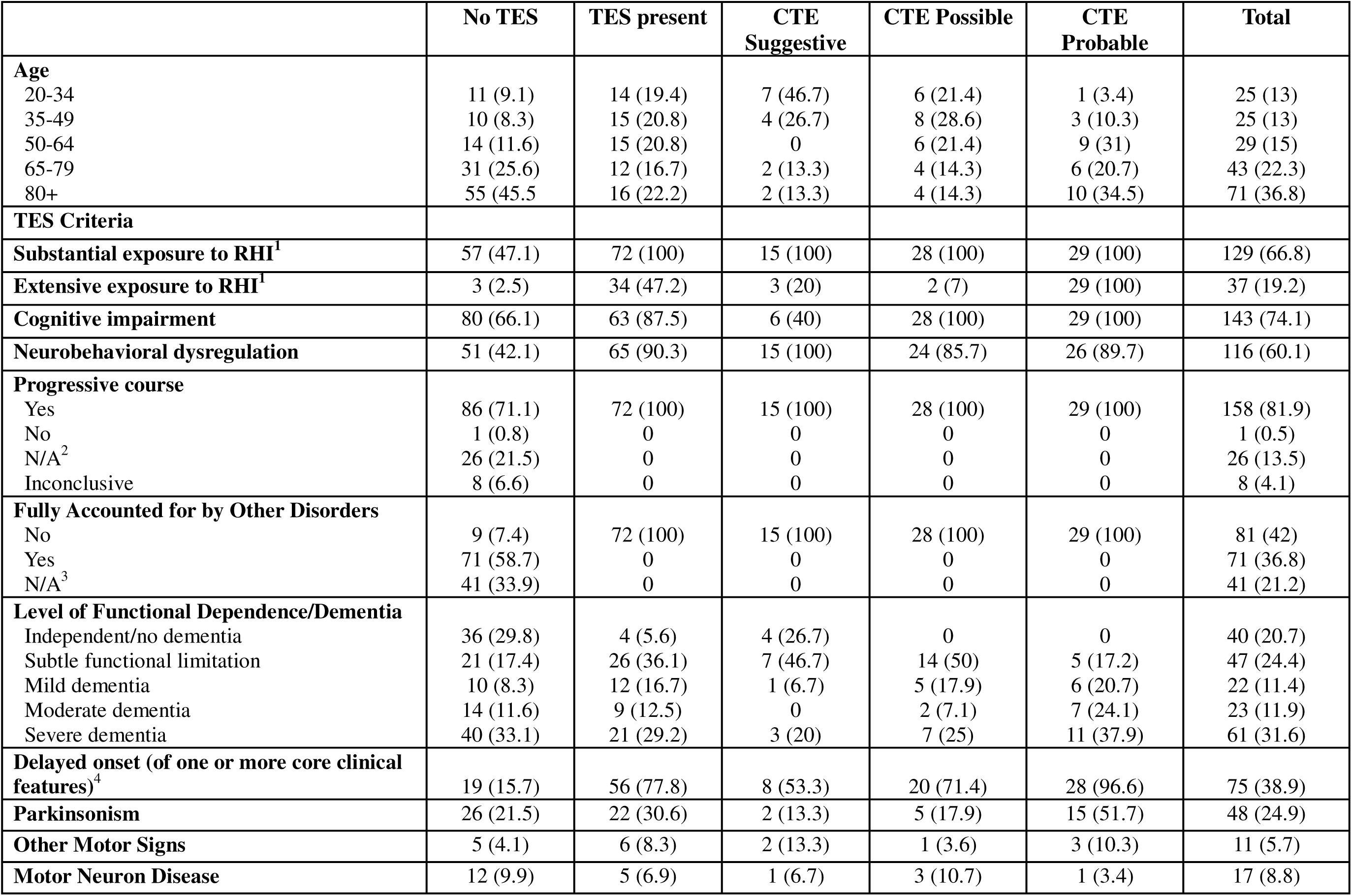
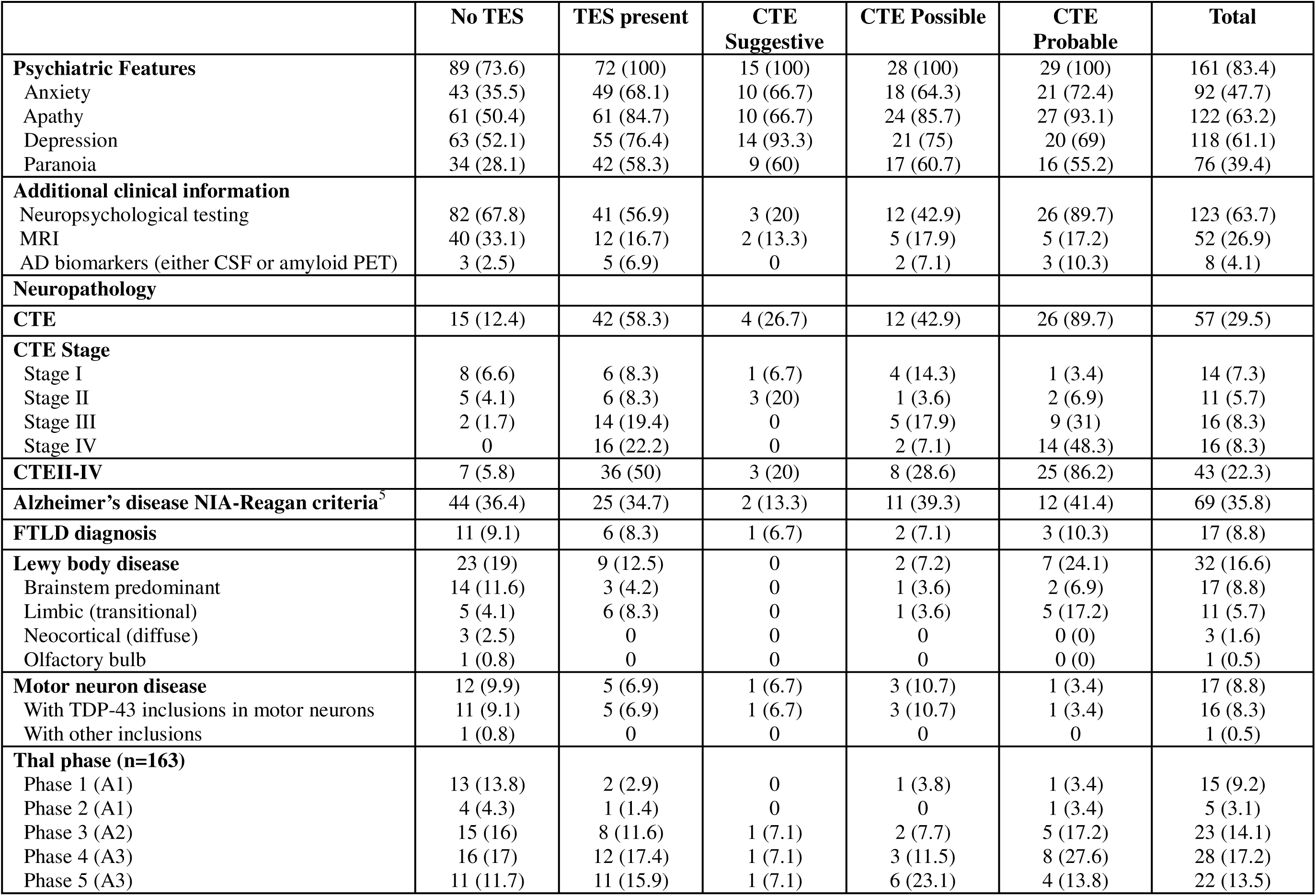
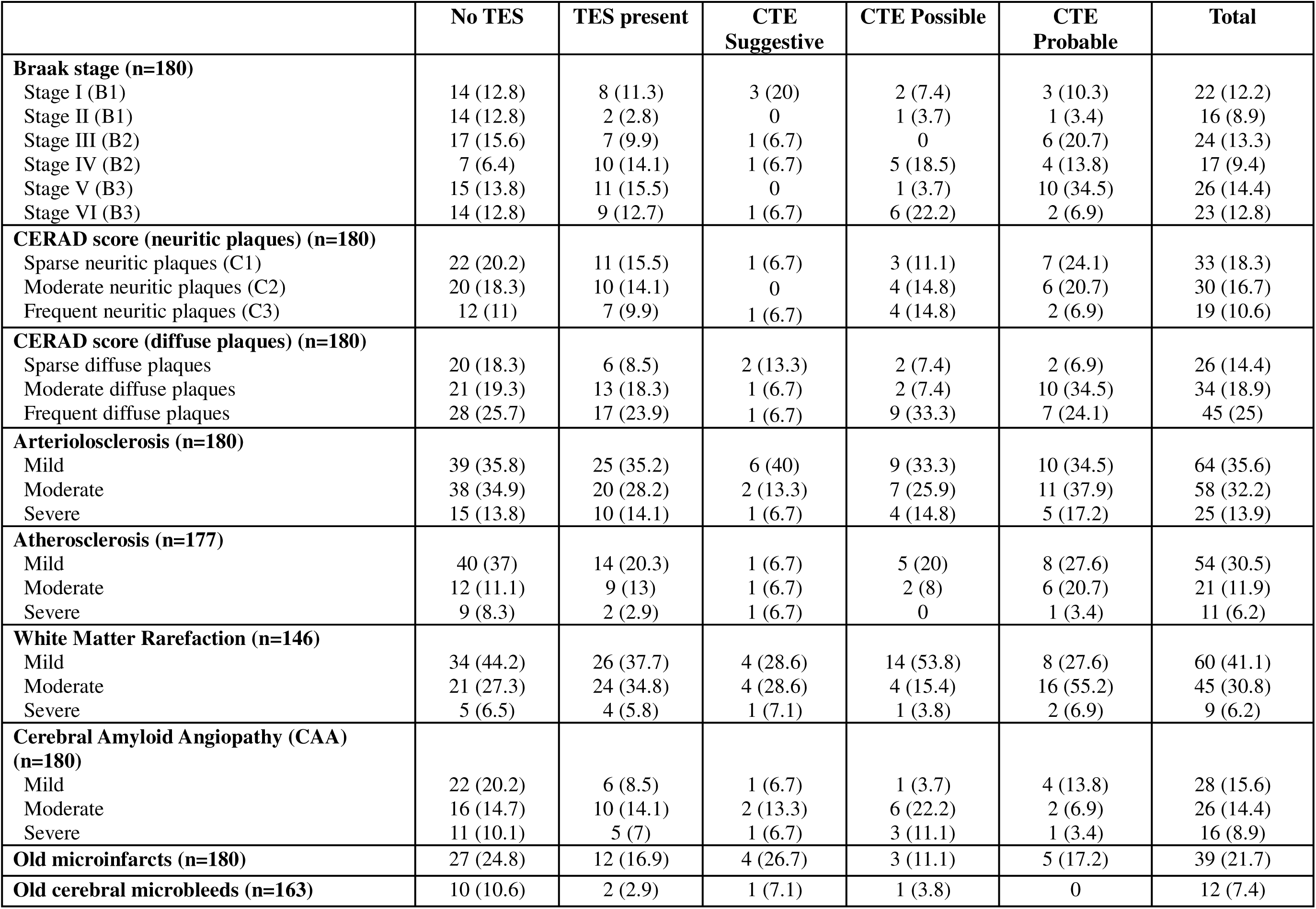
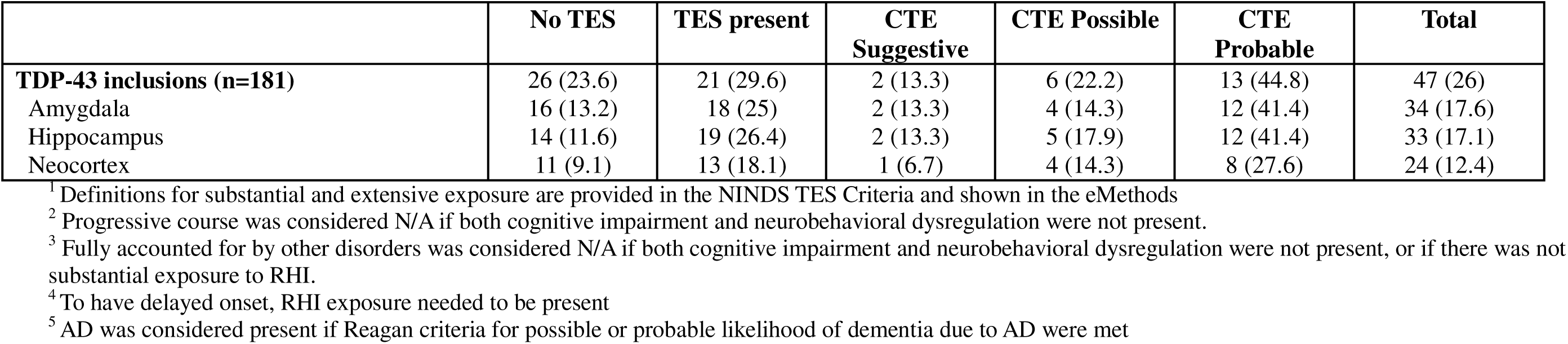
Descriptives and Neuropathology (stratified by TES Status)

Descriptive and neuropathological characteristics are provided in the supplement stratified by six brain banks **(eTable 2**), nine RHI exposure bins (**eTable 3**), and five age bins (**eTable 4**).

Interrater reliability demonstrated very high agreement between voters (ICC: 0.95, 95% CI 0.88-0.97) for the primary analysis (CTE_pos/prob_ vs. no TES or CTE_sug_). There was one outlier voter (i.e., random effect >1.5 x interquartile range), though results did not meaningfully change when the outlier was removed (ICC: 0.95, 95% CI 0.90-0.98). Sensitivity analyses for alternative dichotomizations showed similar interrater reliability: 1) CTE_sug/pos/prob_ vs. no TES: ICC: 0.89, 95% CI 0.87-0.98; 2) CTE_prob_ vs. no TES or CTE_sug/pos_: ICC: 0.97, 95% CI 0.95-0.99.

**Table 2** shows eleven 2x2 tables with TES clinical diagnoses by CTE neuropathological diagnoses. The center 2x2 table shows the primary dichotomizations for clinical determination (CTE_pos/prob_ vs. no TES or CTE_sug_) and neuropathological diagnosis (CTE stages II-IV vs. no CTE or CTE stage I). Additionally, there are eight sensitivity analyses with different dichotomizations for clinical (CTE_sug/pos/prob_ vs. no TES; CTEprob vs. no TES or CTE_sug/pos_) and neuropathological measures (CTE stages I-IV vs. no CTE; CTE stages III-IV vs. no CTE or CTE stages I-II) and two tables showing results stratified by age at death before and after age 50. In the primary analyses, CTE_pos/prob_ demonstrated good sensitivity (0.77, 95%CI:0.64-0.89) and specificity (0.84, 95%CI:0.78-0.90), and good evidence to rule out (LR-: 0.28, 95%CI:0.15-0.50) and to rule in CTE stages II-IV (LR+: 4.8, 95%CI:3.02-7.61). Among donors with age at death ≥50, CTE_pos/prob_ demonstrated very good sensitivity (0.90, 95%CI:0.80-1) and specificity (0.90, 95%CI:0.85-0.96), and strong evidence to rule out (LR-: 0.11, 95%CI:0.04-0.33) and to rule in CTE stages II-IV (LR+: 9.2, 95%CI:4.9-17.27). In sensitivity analyses examining alternative dichotomizations for clinical and pathological diagnoses, specificity and LR+ improved at the expense of worsening sensitivity and LR- as TES thresholds moved up (CTE_sug/pos/prob_ ➔ CTE_pos/prob_ ➔ CTEprob). Sensitivity and LR- worsened without affecting specificity or LR+ with the inclusion of lower stages of pathology (CTE stages III-IV ➔ CTE stages II-IV ➔ CTE stages I-IV). Additional sensitivity analyses (**eTable 5**) showed that performance remained similar when AD pathology was present (n=69; LR+=5.2, 95%CI:2.5-10.7; LR-=0.15, 95%CI:0.04-0.58), when another non-CTE and non-AD neurodegenerative pathology was present (n=57; LR+=8.8, 95%CI:3.5-22.2; LR-=0), and when only non-football playing donors (n=122; LR+=9.2, 95%CI:4.6-18.4; LR-=0.18, 95%CI:0.05-0.69) were included. There were reductions in performance among those with a consensus diagnosis of substance use disorder (n=56; LR+=2.2, 95%CI:0.98-4.9; LR-=0.58, 95%CI:0.27-1.24) and mental health disorder (n=88; LR+=2.9, 95%CI:1.5-5.6; LR-=0.47, 95%CI:0.24-0.94). Performance improved when neuropsychological test scores were available (n=123; LR+=7.6, 95%CI:4.0-14.4; LR-=0.11, 95%CI:0.04-0.35). Analyses using “substantial” or “extensive” RHI exposure levels instead of CTE_pos/prob_ were excellent at ruling out CTE pathology (LR-=0.06, 95%CI:0.01-0.39), but poor at ruling it in (LR+=1.7, 95%CI:1.3-2.2). Analyses using “substantial” or “extensive” RHI exposure levels and the core feature of cognitive impairment, instead of considering all clinical features of CTE_pos/prob,_ demonstrated similar ability to rule out CTE pathology as the full criteria (LR-=0.30, 95%CI:0.15-0.59) but with reduced ability to rule in CTE pathology (LR+=2.1, 95%CI:1.5-3.0) (**eTable 5)**. Analyses using “extensive” RHI exposure instead of CTEprob were excellent, but still not as good at ruling in CTE pathology (LR+=17.4, 95%CI:7.8-39.1), and were better, but not excellent at ruling out CTE pathology (LR-=0.31, 95%CI:0.20-0.50).

**Table 2.**
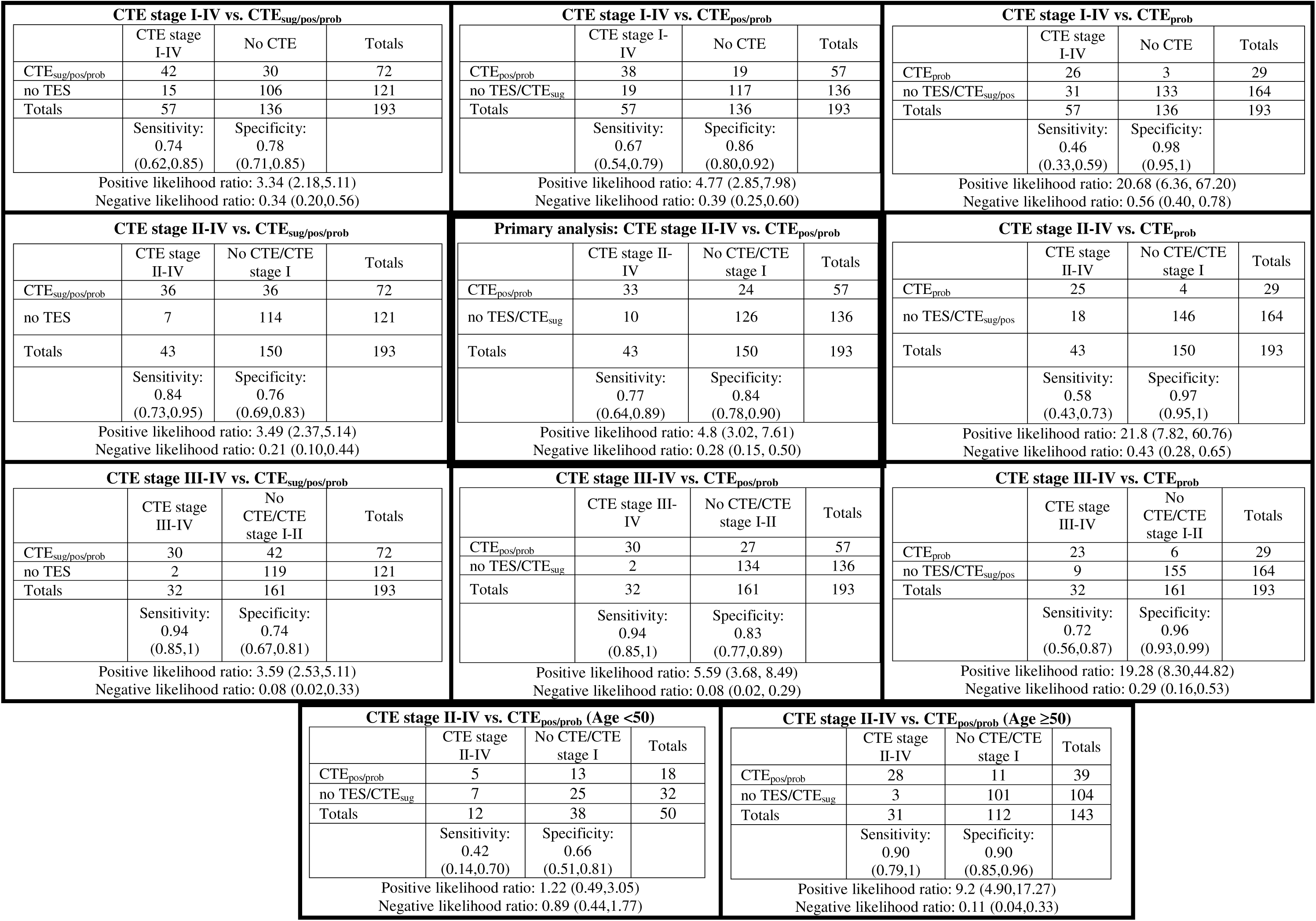
TES consensus diagnoses by CTE pathology.

| CTE stage I-IV vs. CTE<sub>sug/pos/prob</sub> |  |  |  | CTE stage I-IV vs. CTE<sub>pos/prob</sub> |  |  |  | CTE stage I-IV vs. CTE<sub>prob</sub> |  |  |  |
| --- | --- | --- | --- | --- | --- | --- | --- | --- | --- | --- | --- |
|  | CTE stage I-IV | No CTE | Totals |  | CTE stage I-IV | No CTE | Totals |  | CTE stage I-IV | No CTE | Totals |
| CTE<sub>sug/pos/prob</sub> | 42 | 30 | 72 | CTE<sub>pos/prob</sub> | 38 | 19 | 57 | CTE<sub>prob</sub> | 26 | 3 | 29 |
| no TES | 15 | 106 | 121 | no TES/CTE<sub>sug</sub> | 19 | 117 | 136 | no TES/CTE<sub>sug/pos</sub> | 31 | 133 | 164 |
| Totals | 57 | 136 | 193 | Totals | 57 | 136 | 193 | Totals | 57 | 136 | 193 |
|  | Sensitivity: 0.74 (0.62,0.85) | Specificity: 0.78 (0.71,0.85) |  |  | Sensitivity: 0.67 (0.54,0.79) | Specificity: 0.86 (0.80,0.92) |  |  | Sensitivity: 0.46 (0.33,0.59) | Specificity: 0.98 (0.95,1) |  |
| Positive likelihood ratio: 3.34 (2.18,5.11)<br>Negative likelihood ratio: 0.34 (0.20,0.56) |  |  |  | Positive likelihood ratio: 4.77 (2.85,7.98)<br>Negative likelihood ratio: 0.39 (0.25,0.60) |  |  |  | Positive likelihood ratio: 20.68 (6.36, 67.20)<br>Negative likelihood ratio: 0.56 (0.40, 0.78) |  |  |  |
| CTE stage II-IV vs. CTE<sub>sug/pos/prob</sub> |  |  |  | Primary analysis: CTE stage II-IV vs. CTE<sub>pos/prob</sub> |  |  |  | CTE stage II-IV vs. CTE<sub>prob</sub> |  |  |  |
|  | CTE stage II-IV | No CTE/CTE stage I | Totals |  | CTE stage II-IV | No CTE/CTE stage I | Totals |  | CTE stage II-IV | No CTE/CTE stage I | Totals |
| CTE<sub>sug/pos/prob</sub> | 36 | 36 | 72 | CTE<sub>pos/prob</sub> | 33 | 24 | 57 | CTE<sub>prob</sub> | 25 | 4 | 29 |
| no TES | 7 | 114 | 121 | no TES/CTE<sub>sug</sub> | 10 | 126 | 136 | no TES/CTE<sub>sug/pos</sub> | 18 | 146 | 164 |
| Totals | 43 | 150 | 193 | Totals | 43 | 150 | 193 | Totals | 43 | 150 | 193 |
|  | Sensitivity: 0.84 (0.73,0.95) | Specificity: 0.76 (0.69,0.83) |  |  | Sensitivity: 0.77 (0.64,0.89) | Specificity: 0.84 (0.78,0.90) |  |  | Sensitivity: 0.58 (0.43,0.73) | Specificity: 0.97 (0.95,1) |  |
| Positive likelihood ratio: 3.49 (2.37,5.14)<br>Negative likelihood ratio: 0.21 (0.10,0.44) |  |  |  | Positive likelihood ratio: 4.8 (3.02, 7.61)<br>Negative likelihood ratio: 0.28 (0.15, 0.50) |  |  |  | Positive likelihood ratio: 21.8 (7.82, 60.76)<br>Negative likelihood ratio: 0.43 (0.28, 0.65) |  |  |  |
| CTE stage III-IV vs. CTE<sub>sug/pos/prob</sub> |  |  |  | CTE stage III-IV vs. CTE<sub>pos/prob</sub> |  |  |  | CTE stage III-IV vs. CTE<sub>prob</sub> |  |  |  |
|  | CTE stage III-IV | No CTE/CTE stage I-II | Totals |  | CTE stage III-IV | No CTE/CTE stage I-II | Totals |  | CTE stage III-IV | No CTE/CTE stage I-II | Totals |
| CTE<sub>sug/pos/prob</sub> | 30 | 42 | 72 | CTE<sub>pos/prob</sub> | 30 | 27 | 57 | CTE<sub>prob</sub> | 23 | 6 | 29 |
| no TES | 2 | 119 | 121 | no TES/CTE<sub>sug</sub> | 2 | 134 | 136 | no TES/CTE<sub>sug/pos</sub> | 9 | 155 | 164 |
| Totals | 32 | 161 | 193 | Totals | 32 | 161 | 193 | Totals | 32 | 161 | 193 |
|  | Sensitivity: 0.94 (0.85,1) | Specificity: 0.74 (0.67,0.81) |  |  | Sensitivity: 0.94 (0.85,1) | Specificity: 0.83 (0.77,0.89) |  |  | Sensitivity: 0.72 (0.56,0.87) | Specificity: 0.96 (0.93,0.99) |  |
| Positive likelihood ratio: 3.59 (2.53,5.11)<br>Negative likelihood ratio: 0.08 (0.02,0.33) |  |  |  | Positive likelihood ratio: 5.59 (3.68, 8.49)<br>Negative likelihood ratio: 0.08 (0.02, 0.29) |  |  |  | Positive likelihood ratio: 19.28 (8.30,44.82)<br>Negative likelihood ratio: 0.29 (0.16,0.53) |  |  |  |
| CTE stage II-IV vs. CTE<sub>pos/prob</sub> (Age <50) |  |  |  | CTE stage II-IV vs. CTE<sub>pos/prob</sub> (Age ≥50) |  |  |  |  |  |  |  |
|  | CTE stage II-IV | No CTE/CTE stage I | Totals |  | CTE stage II-IV | No CTE/CTE stage I | Totals |  |  |  |  |
| CTE<sub>pos/prob</sub> | 5 | 13 | 18 | CTE<sub>pos/prob</sub> | 28 | 11 | 39 |  |  |  |  |
| no TES/CTE<sub>sug</sub> | 7 | 25 | 32 | no TES/CTE<sub>sug</sub> | 3 | 101 | 104 |  |  |  |  |
| Totals | 12 | 38 | 50 | Totals | 31 | 112 | 143 |  |  |  |  |
|  | Sensitivity: 0.42 (0.14,0.70) | Specificity: 0.66 (0.51,0.81) |  |  | Sensitivity: 0.90 (0.79,1) | Specificity: 0.90 (0.85,0.96) |  |  |  |  |  |
| Positive likelihood ratio: 1.22 (0.49,3.05)<br>Negative likelihood ratio: 0.89 (0.44,1.77) |  |  |  | Positive likelihood ratio: 9.2 (4.90,17.27)<br>Negative likelihood ratio: 0.11 (0.04,0.33) |  |  |  |  |  |  |  |

Figure 1 displays individual donor-level data, including item-level components of the TES criteria, non-TES clinician consensus diagnoses, and neuropathological features of the 24 false positives (those with a determination of CTE_pos/prob_ without CTE stages II-IV, fig. 1a) and 10 false negatives (those without a determination of CTE_pos/prob_ with CTE stages II-IV, fig. 1b). All false positives under the age of 50 were also diagnosed by the clinician consensus panel with a mental health disorder, substance use disorder and/or a pain disorder. All false positives above the age of 50 had at least one non-CTE neurodegenerative pathological diagnosis, along with substantial vascular pathological burden. Among the 10 false negatives, 7 (70%) were below age 50 and 8 (80%) had stage II CTE. Clinically, 3 (30%) were diagnosed with CTE_sug_, 2 (20%) lacked both core features of cognitive impairment and neurobehavioral dysregulation, 3 (30%) had another etiology fully accounting for core features (Alzheimer’s and vascular dementia, schizophrenia, bipolar disorder with psychotic features), 1 (10%) lacked a progressive course among the core features, and 1 (10%) had insufficient RHI exposure.

**Figure 1:**
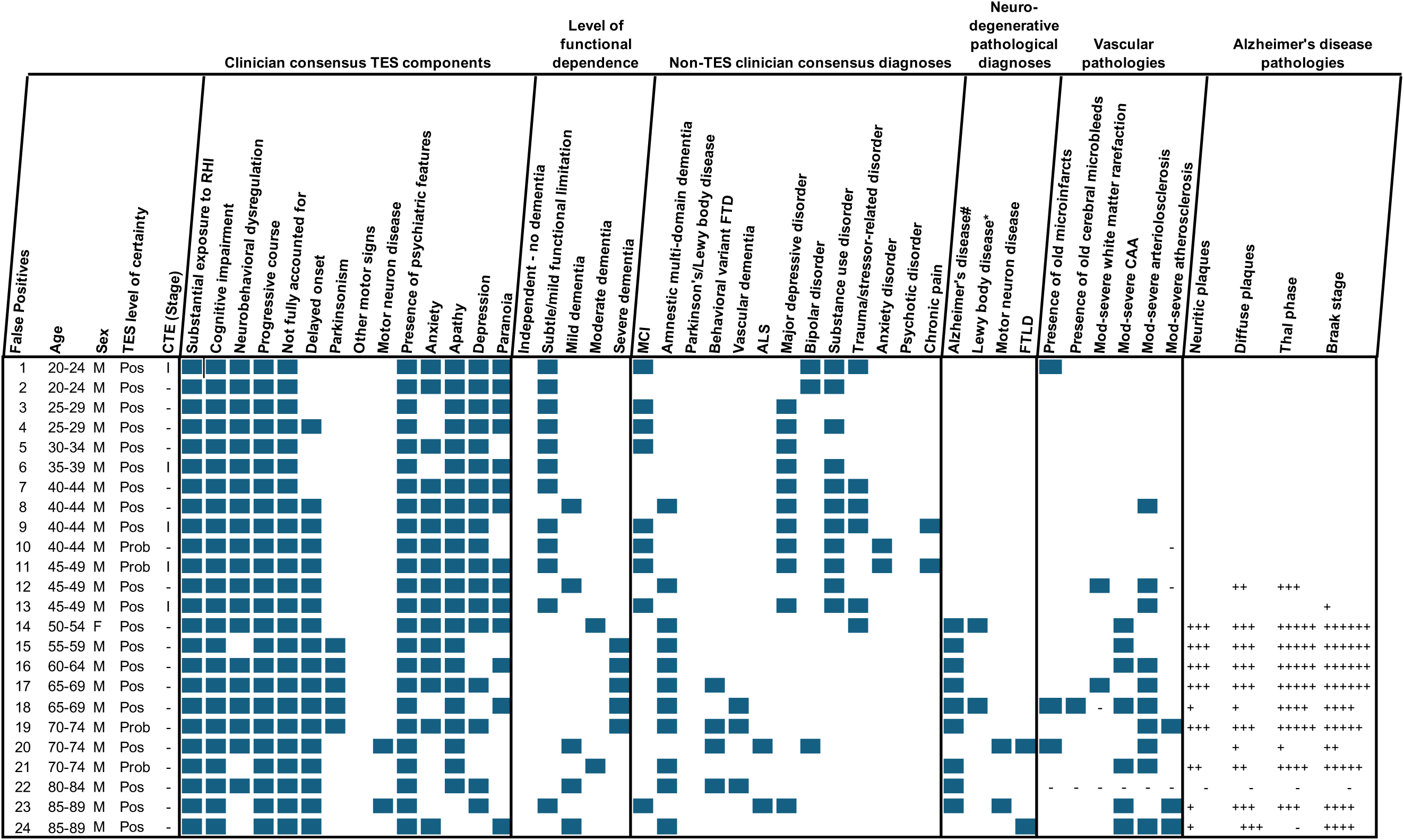
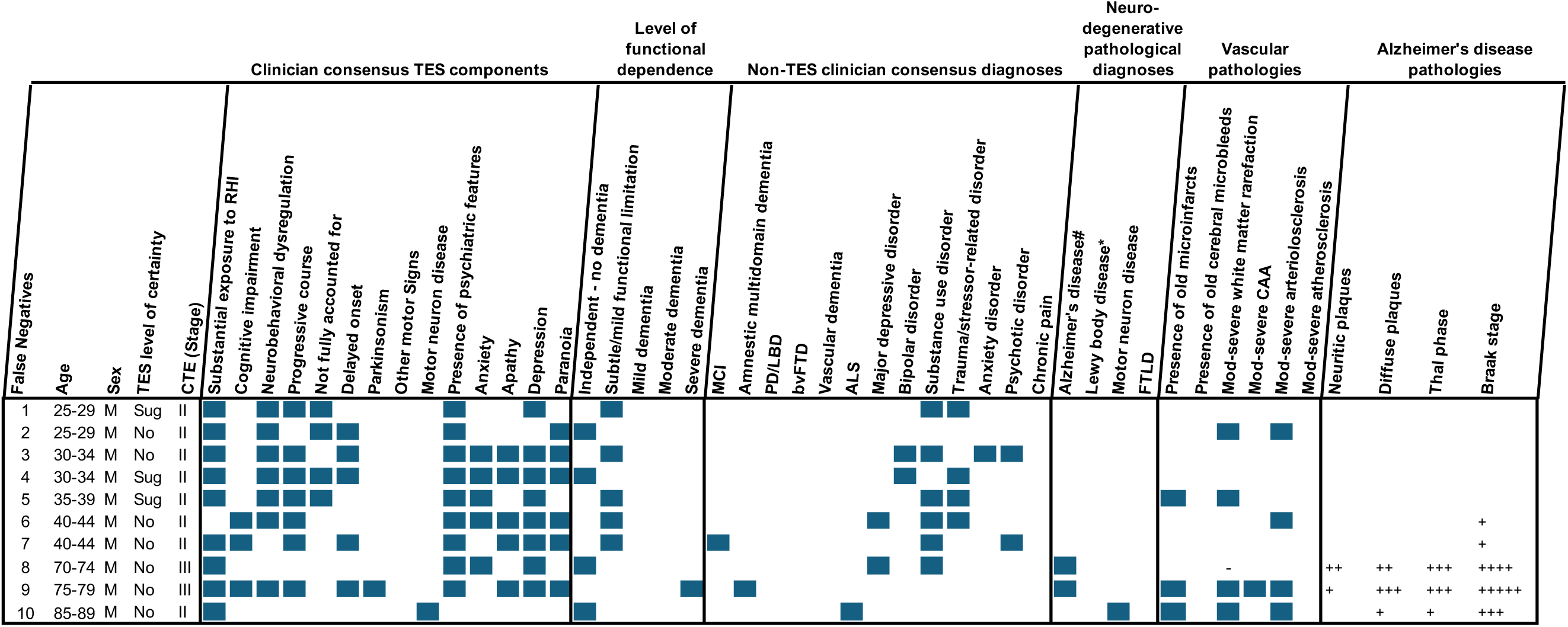
False Positives and Negatives. Individual donor-level data, including item-level components of the TES criteria, non-TES clinician consensus diagnoses, and neuropathological features of the A) 24 false positives (those with a determination of CTE_pos/prob_ without CTEII-IV) and B) 10 false negatives (those without a determination of CTE_pos/prob_ with CTEII-IV). #Alzheimer’s disease (AD) pathology was considered present if Reagan criteria for possible or probable likelihood of dementia due to AD was present. *Lewy body disease pathology was considered present if pathology reached limbic or neocortical levels.

**Table 3** shows item level components of the TES criteria, stratified by levels of RHI exposure (as defined in the NINDS TES Criteria, see **eMethods**) and CTE stage II-IV status.

**Table 3.** TES criteria, stratified by exposure level and CTE status.

|  | All Participants |  | None/Less than Substantial Exposure<sup>1</sup> |  | Substantial Exposure<sup>1</sup> |  |  |  |
| --- | --- | --- | --- | --- | --- | --- | --- | --- |
|  |  |  |  |  | Not Extensive Exposure<sup>1</sup> |  | Extensive Exposure<sup>1</sup> |  |
|  | No CTE or CTE I | CTEII-IV | No CTE or CTE I | CTEII-IV | No CTE or CTE I | CTEII-IV | No CTE or CTE I | CTEII-IV |
| **Number (%)** | 150 | 43 | 63 | 1 | 81 | 12 | 6 | 30 |
| **Age at death (years), SD** | 67.7 (22.7) | 61.8 (18.8) | 82.1 (12.6) | 42.0 (n/a) | 57.8 (22.8) | 64.1(16.7) | 48.7 (20.8) | 61.5 (19.7) |
| **TES status** |  |  |  |  |  |  |  |  |
| No | 114 (76) | 7 (16.3) | 63 (100) | 1 (100) | 50 (61.7) | 4 (33) | 1 (17) | 2 (7) |
| Suggestive | 12 (8) | 3 (7) | 0 | 0 | 11 (13.6) | 1 (8.3) | 1 (17) | 2 (7) |
| Possible | 20 (13.3) | 8 (18.6) | 0 | 0 | 20 (24.7) | 6 (50) | 0 | 2 (7) |
| Probable | 4 (2.7) | 25 (58.1) | 0 | 0 | 0 | 1 (8.3) | 4 (67) | 24 (80) |
| **Cognitive impairment** | 107 (71.3) | 36 (83.7) | 50 (79.4) | 1 (100) | 53 (65.4) | 9 (75) | 4 (67) | 26 (87) |
| **Neurobehavioral dysregulation** | 79 (52.7) | 37 (86) | 17 (27) | 1 (100) | 57 (70.4) | 8 (67) | 5 (83) | 28 (93) |
| **Progressive course** |  |  |  |  |  |  |  |  |
| Yes | 118 (78.7) | 40 (93) | 50 (79.4) | 1 (100) | 62 (76.5) | 10 (83.3) | 6 (100) | 29 (97) |
| No | 1 (0.7) | 0 | 0 | 0 | 1 (1.2) | 0 | 0 | 0 |
| N/A<sup>2</sup> | 24 (16) | 2 (4.7) | 13 (20.6) | 0 | 11 (13.6) | 2 (16.7) | 0 | 0 |
| Inconclusive | 7 (4.7) | 1 (2.3) | 0 | 0 | 7 (8.6) | 0 | 0 | 1 (3) |
| **Fully Accounted for by Other Disorders** |  |  |  |  |  |  |  |  |
| No | 44 (29.3) | 37 (86) | 4 (6.3) | 0 | 35 (43.2) | 8 (67) | 5 (83) | 29 (97) |
| Yes | 68 (45.3) | 3 (7) | 32 (50.8) | 0 | 35 (43.2) | 2 (16.7) | 1 (17) | 1 (3) |
| N/A<sup>3</sup> | 38 (25.3) | 3 (7) | 27 (42.9) | 1 (100) | 11 (13.6) | 2 (16.7) | 0 | 0 |
| **Level of Functional Dependence/Dementia** |  |  |  |  |  |  |  |  |
| Independent/no dementia | 36 (24) | 4 (9.3) | 12 (19) | 0 | 23 (28.4) | 2 (16.7) | 1 (17) | 2 (7) |
| Subtle functional limitation | 35 (23.3) | 12 (27.9) | 4 (6.3) | 1 (33) | 28 (34.6) | 4 (33.3) | 3 (50) | 7 (23) |
| Mild dementia | 16 (10.7) | 6 (14) | 4 (6.3) | 0 | 12 (14.8) | 0 | 0 | 6 (20) |
| Moderate dementia | 16 (10.7) | 7 (16.3) | 10 (15.9) | 0 | 5 (6.2) | 2 (16.7) | 1 (17) | 5 (17) |
| Severe dementia | 47 (31.3) | 14 (32.6) | 33 (52.4) | 0 | 13 (16) | 4 (33.3) | 1 (17) | 10 (33) |
| **Delayed onset (of one or more core clinical features)<sup>4</sup>** | 40 (26.7) | 35 (81.4) | 1 (1.6) | 0 | 35 (43.2) | 9 (75) | 4 (67) | 26 (87) |
|  |  |  |  |  | Not Extensive Exposure<sup>1</sup> |  | Extensive Exposure<sup>1</sup> |  |
|  | No CTE or CTE I | CTEII-IV | No CTE or CTE I | CTEII-IV | No CTE or CTE I | CTEII-IV | No CTE or CTE I | CTEII-IV |
| **Parkinsonism** | 32 (21.3) | 16 (37.2) | 14 (22.2) | 0 | 17 (21) | 2 (16.7) | 1 (17) | 14 (47) |
| **Other Motor Signs** | 7 (4.7) | 4 (9.3) | 5 (7.9) | 0 | 2 (2.5) | 2 (16.7) | 0 | 2 (7) |
| **Motor Neuron Disease** | 14 (9.3) | 3 (7) | 5 (7.9) | 0 | 9 (11.1) | 2 (16.7) | 0 | 1 (3) |
| **Psychiatric Features** | 119 (79.3) | 42 (97.7) | 46 (73) | 1 (100) | 67 (82.7) | 11 (91.7) | 6 (100) | 30 (100) |
| Anxiety | 64 (42.7) | 28 (65.1) | 16 (25.4) | 1 (100) | 44 (54.3) | 5 (41.7) | 4 (67) | 22 (73) |
| Apathy | 88 (58.7) | 34 (79.1) | 38 (60.3) | 1 (100) | 45 (55.6) | 8 (67) | 5 (83) | 25 (83) |
| Depression | 87 (58) | 31 (72.1) | 26 (41.3) | 1 (100) | 57 (70.4) | 8 (67) | 4 (67) | 22 (73) |
| Paranoia | 51 (34) | 25 (58.1) | 11 (17.5) | 1 (100) | 37 (45.7) | 6 (50) | 3 (50) | 18 (60) |

<sup>1</sup> Definitions for substantial and extensive exposure are provided in the NINDS TES Criteria and shown in the eMethods
<sup>2</sup> Progressive course was considered N/A if both cognitive impairment and neurobehavioral dysregulation were not present.
<sup>3</sup> Fully accounted for by other disorders was considered N/A if both cognitive impairment and neurobehavioral dysregulation were not present, or if there was not substantial exposure to RHI.
<sup>4</sup> To have delayed onset, RHI exposure needed to be present.

Nearly all donors with less than substantial RHI exposure did not have CTE stages II-IV (63/64, 98%). Donors with substantial, but not extensive exposure had CTE stages II-IV inconsistently (12/93, 13%). Donors with extensive RHI exposure had CTE stages II-IV frequently (30/36, 83%). Across all donors, cognitive impairment (36/43, 83.7% vs. 107/150, 71.3%), neurobehavioral dysregulation (37/43, 86% vs. 79/150, 52.7%) and a progressive course (40/43, 93%, 118/150, 78.7%) were more frequently identified in those with CTE stages II-IV than those without. Other supportive features found more frequently in those with CTE stages II-IV than those without included dementia (27/43, 62.8% vs. 79/150, 52.7%) a delayed onset of core clinical features (35/43, 81.4% vs. 40/150, 26.7%), parkinsonism (16/43, 37.2% vs. 32/150, 21.3%), and psychiatric features (42/43, 98% vs. 119/150, 79.3%). Item level components of the TES criteria stratified by CTE stage are presented in **eTable 6.**

Figure 2 displays three Venn diagrams of TES item level data among donors with A) both cognitive impairment and neurobehavioral dysregulation (92/193, 47.7%), B) cognitive impairment, but not neurobehavioral dysregulation (51/193, 26.4%) and C) neurobehavioral dysregulation, but not cognitive impairment (24/193, 12.4%). Among cases with CTE stages II-IV, most fell into A) having both cognitive impairment and neurobehavioral dysregulation [32/43 (74.4%)]. Across all 3 diagrams, among cases with CTE stages II-IV, most were in the center having substantial RHI exposure, a progressive course and being not fully accounted for by another process [36/43 (83.7%)].

**Figure 2:**
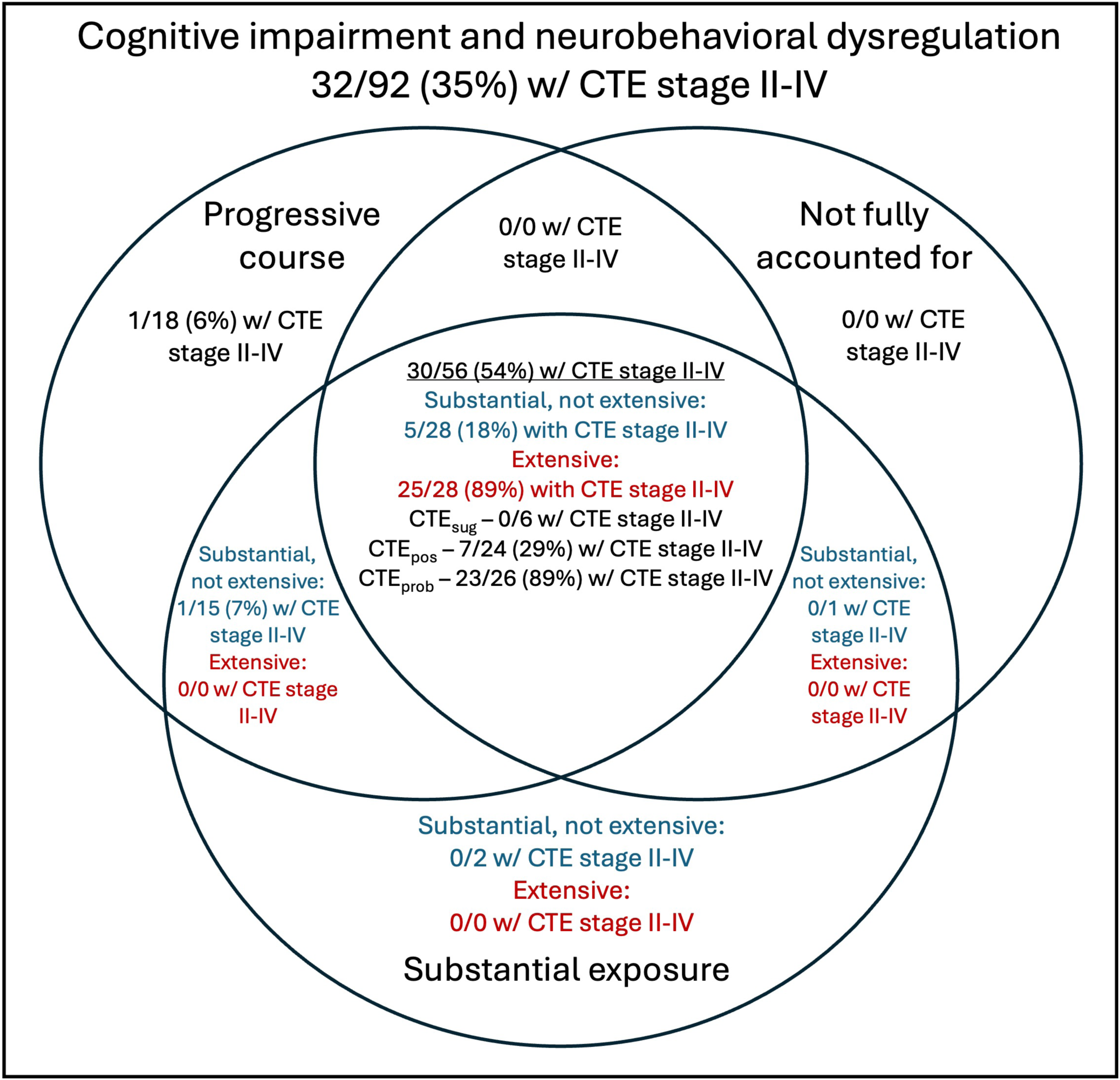
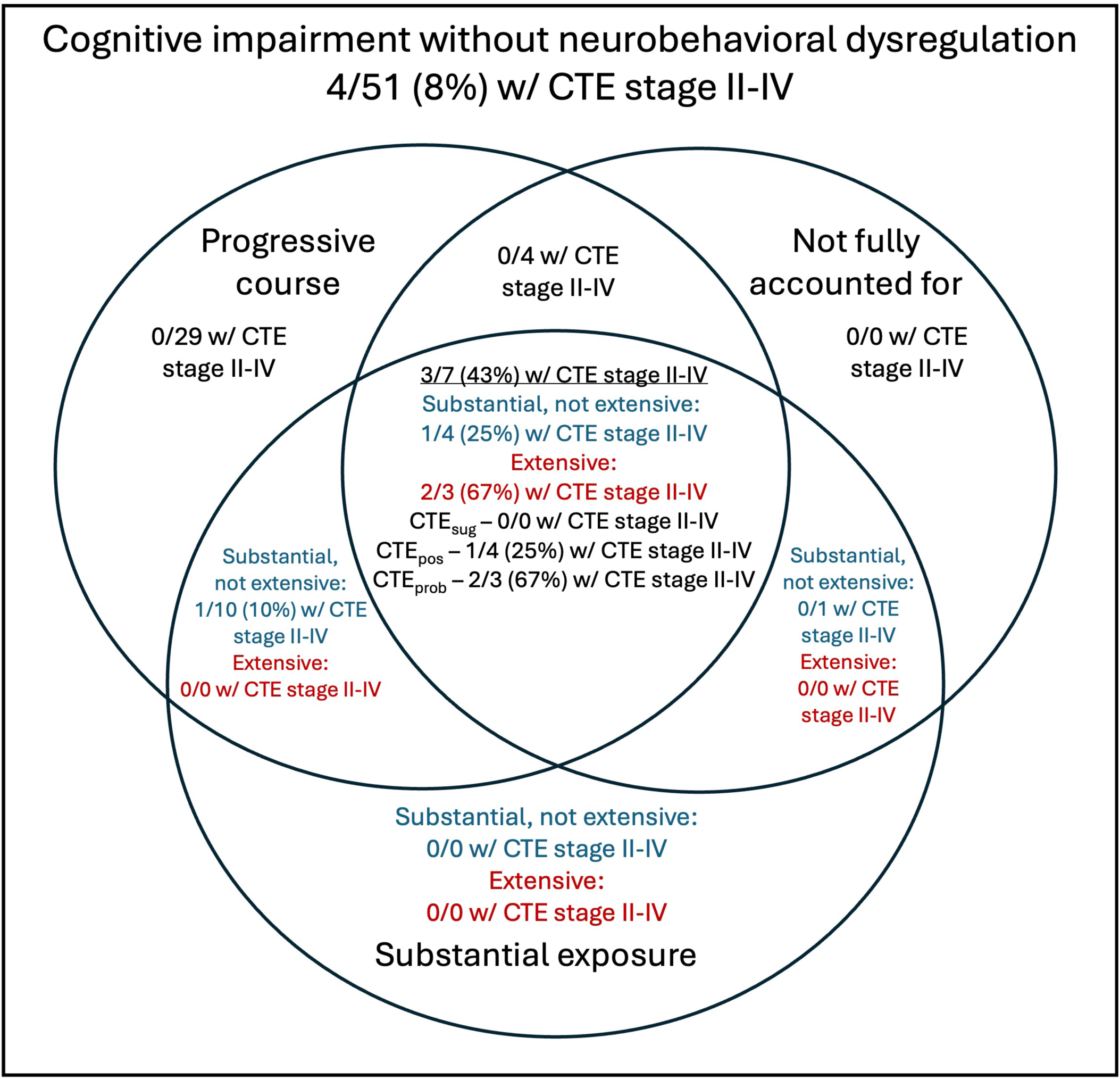
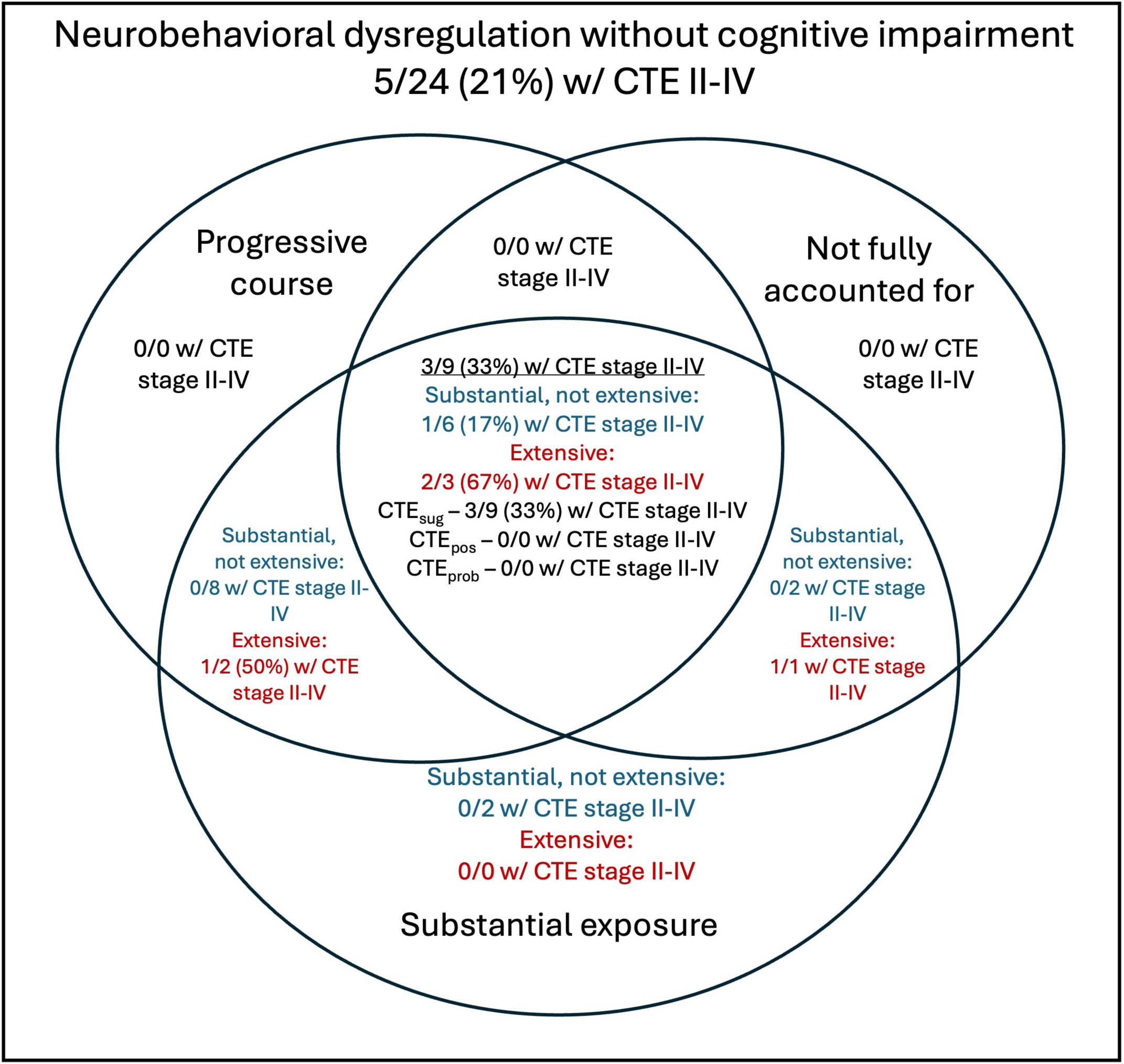
**Venn diagrams of TES item level data** among donors with A) both cognitive impairment and neurobehavioral dysregulation, B) cognitive impairment, but not neurobehavioral dysregulation and C) neurobehavioral dysregulation, but not cognitive impairment. Each circle is a required criterion for an NINDS TES determination and only the central overlap represents donors with a positive diagnosis. Definitions for substantial and extensive exposure are provided in the NINDS TES Criteria and shown in the eMethods. Progressive course was considered N/A if both cognitive impairment and neurobehavioral dysregulation were not present. No fully accounted for by other disorders was considered N/A if both cognitive impairment and neurobehavioral dysregulation were not present, or if there was not substantial exposure to RHI.

Among donors with substantial RHI exposure, the accuracy of CTE_pos/prob_ to predict CTE stages II-IV did not significantly differ when AD pathology was present vs. absent (70% vs.76%, p=0.50).

## Discussion

In this study we assessed the interrater reliability and validity of the NINDS proposed research diagnostic criteria for TES, using CTE pathology as the gold standard. Among 193 brain donors recruited across a range of ages, RHI exposures and brain banks, the NINDS criteria demonstrated good reliability, sensitivity and specificity, and provided moderate to large evidence to both rule out and rule in CTE pathology. Among donors older than 50, as well as among those with neuropsychological data available, the criteria performance was further improved. False positive cases under age 50 commonly had substance use, psychiatric and/or chronic pain diagnoses, while older false positive cases had non-CTE neurodegenerative and vascular pathologies present. Most false negatives were young and had stage II CTE. Nearly all cases with CTE had substantial RHI exposure, and most had both core clinical features of cognitive impairment and neurobehavioral dysregulation.

The 2014 TES criteria were developed to prioritize sensitivity with the tradeoff of low specificity, similar to early criteria for other neurodegenerative diseases.^23,49^ Assessment of the 2014 criteria among RHI-exposed brain donors, indeed showed high sensitivity at the expense of low specificity. The NINDS criteria, in turn, were tailored to improve specificity while preserving sensitivity.^26^ Results from the current study indeed demonstrate improved specificity (0.21 to 0.84) with an acceptable reduction in sensitivity (0.97 to 0.77). Specificity and LR+ increased with each successive provisional level for CTE certainty (CTE_sug/pos/prob_ ➔ CTE_pos/prob_ ➔ CTE_prob_).^26^ Sensitivity and specificity for CTE_pos/prob_ were comparable to recent AD, FTD, and DLB criteria.^50–53^

Our findings provide insight into the settings when the TES criteria are most valid and when they could be improved. Specificity, sensitivity, LR+ and LR- were all compelling among donors over age 50 and those with neuropsychological testing. The criteria may perform better in older individuals due to the presence of more extensive CTE pathology, which is associated with age, more severe cognitive symptoms and dementia.^7,16^ Neuropsychological test scores provide standardized, objective evidence of cognitive impairment, as opposed to subjective report of cognitive symptoms from a patient or informant. Indeed, the NINDS criteria state that cognitive impairment should be substantiated by impaired performance on neuropsychological testing if available,^26^ and diagnostic criteria for other neurodegenerative diseases require cognitive impairment to be validated by neuropsychological testing.^50,54–56^ Interestingly, other settings in which we suspected the criteria to perform better, including absence of AD pathology (due to overlapping clinical features between AD and CTE as well as performance of the 2014 criteria) and among football players (where exposure cut points are most clearly specified), did not show improved performance.

As in the study of the 2014 criteria^23^, the current study had more false positives than false negatives, although the gap was comparatively reduced. All false positives under age 50 were also diagnosed by the clinician consensus panel with mental health disorders, substance use disorder, and/or chronic pain that may have led to some of the symptoms required for a TES determination. In these cases, the consensus panel did not *fully* attribute the clinical syndrome to these alternative diagnoses, a substantial bar to reach. Consistently, both sensitivity and specificity were reduced in the presence of substance use and mental health disorders. In the 2014 study, each of the eight false positives with no other neurodegenerative or cerebrovascular pathology likewise were diagnosed with a mental health disorder, substance use disorder, and/or chronic pain.^23^ These findings highlight the complexity of identifying the underlying etiology of neuropsychiatric symptoms among young individuals with a history of RHI. Further, the workup of neurodegenerative disease requires identifying major psychiatric disorders and addressing treatable conditions^57^ and these findings underscore the importance of these steps when considering a TES determination.

Conversely, false positives over age 50 largely did not have psychiatric or substance use disorders, but instead had non-CTE neurodegenerative pathologies, including AD, LBD, MND, FTLD-tau, and/or FTLD-TDP. However, in sensitivity analyses only among those with AD pathology or only among those with additional non-AD and non-CTE pathology, sensitivity and specificity of the TES criteria were similar to the full sample. Further, among donors with substantial RHI exposure, the accuracy of a TES determination did not significantly differ when AD pathology was present vs. absent.

Most false negatives were younger than age 50 and diagnosed with stage II CTE. For these donors, the relatively low level of CTE pathology was likely insufficient to produce clinical features necessary to meet CTE_pos/prob_ criteria. Indeed, only three met the required cognitive impairment core clinical feature and only one had dementia. An additional two did not have either core clinical feature. A pre-clinical stage during which pathology is present but symptoms have not manifested is a common feature of neurodegenerative diseases,^5,58,59^ and at this time little is known about the pre-clinical period during which symptoms related to CTE neuropathology have not yet manifested. As with the young false positives, the young false negatives had extensive neuropsychiatric features which complicated diagnosis. In three false negatives, the clinician panel fully attributed the syndrome to a mental health disorder. Future iterations of the TES criteria may benefit from a more nuanced approach to the contribution of comorbidities.

We conducted additional sensitivity analyses and explored individual components of the criteria descriptively to see which provided the most utility in predicting CTE. Substantial exposure to RHI clearly had a prominent role, as all but one donor with CTE pathology met this criterion (98%), while 58% of donors without CTE met the criterion. The only donor with CTE stages II-IV who was not determined to have substantial exposure to RHI still had over two decades of exposure to snowboarding, which the clinician panelists did not automatically judge to meet criteria for substantial exposure. Since the NINDS Criteria were proposed in 2021, new years-of-play thresholds have been proposed for ice hockey^3^ and investigations into other sports are currently underway. Although the exposure thresholds are central to the criteria, the additional components of the criteria still added value. Compared to the primary analyses using CTE_pos/prob,_ sensitivity analyses that only used “substantial” or “extensive” RHI exposure thresholds without any clinical features were poor at ruling in CTE, though excellent at ruling it out. Sensitivity analyses that used both “substantial” or “extensive” RHI exposure thresholds and the core feature of cognitive impairment, were better at ruling in CTE, but still not as good as the full criteria. Additionally, presence of both core clinical features together with the other primary diagnostic components in aggregate captured the majority of CTE cases. This may suggest that beyond RHI exposure, the combination of components, rather than any single component, factored into the criteria’s performance.

Recently, Arena et al. investigated the validity of the NINDS TES criteria in the University of Pennsylvania Center for Neurodegenerative Disease Research brain bank where CTE prevalence was very low (1.3%) and claimed that the performance of the criteria was poor because the positive predictive value (PPV) was low (24%).^60^ Importantly, PPV is a function of disease prevalence and invariably falls with falling prevalence. It is for this reason, that we do not report positive and negative predictive values within this manuscript, but instead report positive and negative likelihood ratios, that are independent of prevalence.^61^ Using data from Arena et al., we calculated the LR+ to be 24.3, providing similar, if not better, evidence for the criteria’s ability to rule in CTE pathology. Additionally, Arena et al. did not have access to years of play and instead used particularly liberal proxies (high school play alone, military boxing, pair skating) as sufficient to meet the substantial exposure to RHI criterion (which requires at least 5 years of play for American football and at least as much for other contact sports). Additionally, they considered domestic violence alone to meet the criterion, even though the largest study to date did not find any cases of CTE among 84 brain donors who experienced intimate partner violence.^62^ Excluding these cases would have removed 17 of 25 donors considered to meet the NINDS TES criteria (14 of which were false positives). In contrast, the current study carefully captured all information necessary to assess the substantial exposure to RHI criterion and primary analyses only considered TES with possible or probable levels of CTE certainty, that require elements that have been most clearly linked to CTE pathology (contact sports and cognitive impairment).^26^

Strengths of this study include a diverse sample including five age groups spanning ages 20-106, nine head impact exposure groups, and recruitment from six brain banks, a rigorous study design involving blinded clinicians, neuropathologists and informants, and the participation of a multi-disciplinary panel of expert clinician-voters representing multiple institutions. In contrast with the prior study of the 2014 criteria that exclusively included RHI-exposed brain donors,^23^ this sample was purposefully selected to include varied head impact exposures, including TBI without RHI and no reported head trauma, to broaden generalizability. Limitations of this study include retrospective clinical diagnosis. Clinicians were privy to the entire disease course until death as opposed to a typical clinical setting in which initial diagnosis may occur early in the disease course, when the longitudinal trajectory is less established.

Further, several of the clinicians were involved in developing the NINDS TES criteria and were therefore better versed in applying them than most doctors in the community. Additionally, available data on donors was inconsistent, with only a subset of donors having prospective research assessment in life (n=94, 49%), neuropsychological testing (n=123; 64%) or imaging and fluid biomarkers (n=43; 22%). Indeed, criteria performance was better when neuropsychological testing data were available. Informants may have not been present for a loved one’s athletic career, other RHI or TBI exposure or some of their disease course. However, this lack of consistent data would reduce the performance of the criteria and is closer to a real-world setting, adding further credence to the criteria’s performance. Although our access to six brain banks and careful selection criteria allowed for inclusion of a wide spectrum of exposures and ages, the different demographic and exposure makeups of the brain banks did not allow for demographic and exposure matching across brain banks. Nonetheless, donors with substantial RHI exposure spanned 5 of 6 brain banks (FHS: 17 (53%); BU ADRC: 5 (20%); VA ALS:12 (75%); LETBI: 12 (67%); UNITE: 83 (97%)]. Lastly, to maximize generalizability across age, head impact types and community vs. clinic-based settings, we artificially created a setting that is unlike most specialized clinics that would apply these criteria. For instance, a memory clinic would see older patients with a lower burden of head trauma and a sports concussion clinic would see younger patients, few if any of whom would have dementia. As such, the CTE prevalence in those clinics is unlikely to match the prevalence in our study and we therefore do not report positive and negative predictive values, which are a function of prevalence.

## Conclusions

The NINDS TES criteria demonstrated good reliability, sensitivity and specificity, and provided moderate to large evidence to both rule out and rule in CTE pathology, particularly above age 50 and when neuropsychological testing was present.

## Supporting information

Supplement packet

## Data Availability

Data available:Yes
Data types:Deidentified participant data
Data dictionary
How to access data:Deidentified data can be accessed at FITBIR: https://fitbir.nih.gov/
When available:With publication
Who can access the data:Data will be released to investigators who submit an approved data request to FITBIR.
Types of analyses:Please see the data access criteria on the FITBIR website https://fitbir.nih.gov/

## Acknowledgements

We gratefully acknowledge the help of all members of the Boston University CTE Center, UNITE Brain Bank, Framingham Heart Study, Boston University Alzheimer’s Disease Research Center, Mt. Sinai Alzheimer’s Disease Research Center, VA ALS Biorepository, Late Effects of Traumatic Brain Injury (LETBI) Study and the Concussion & CTE Foundation, as well as the individuals and families whose participation and contributions made this work possible.

## Author Contributions

Mr. Abdolmohammadi, Dr. Durape, Dr. Daneshvar and Dr. Mez had full access to all the data in the study and take responsibility for the integrity of the data and the accuracy of the data analysis. JM and DHD were responsible for study conception and design. JM, BA, SD, and DHD did the analysis and interpreted the results in collaboration with all remaining authors. JM, BA, BD, MLA, BY, AP, NBF, AH, MR, FF, SN, BM, JP, KDOC, LEG, DIK, RCC, NWK, RAS,

VEA, BRH, JFC, TDS, ACM, DHD were responsible for data acquisition. All authors critically revised the paper for important intellectual content and approved the final version.

## Funding

National Institute of Neurological Disorders and Stroke (U54NS115266, U01NS086659, U01NS093334, R01NS078337, R56NS078337, RF1NS143015, R01NS142076, R01NS122854, RF1NS115268, R01NS128961), National Institute on Aging (P30AG072978, P30AG066514, U19AG068753, R01AG061028, K23AG046377, R21HD089088, F32NS096803, RF1AG062348, R01AG077588), National Heart, Lung and Blood Institute (75N92019D00031), Department of Veterans Affairs (I01 CX001135, CSP 501, B6796-C), Department of Defense (W81XWH-13-2-0095, W81XWH-13-2-0064, W81XWH1810580, PRARP-13267017), the Alzheimer’s Association (NIRG-15-362697, NIRG-305779), the National Operating Committee on Standards for Athletic Equipment (NOCSAE), the Nick and Lynn Buoniconti Foundation, the Concussion & CTE Foundation, and a gift from William Collatos.

## Role of the Funder/Sponsor

The funders had no role in the design and conduct of the study; collection, management, analysis, and interpretation of the data; preparation, review, or approval of the manuscript; and decision to submit the manuscript for publication.

## Conflict of Interest Disclosures

Dr Mez reported receiving grants from the NIH outside the submitted work.

Dr. Alosco receives honorarium from the Michael J Fox Foundation for services unrelated to this study. He receives royalties from Oxford University Press Inc. He receives research support from Life Molecular Imaging Inc.

Dr Nowinski reported serving as a volunteer member of the Mackey-White Committee of the National Football League (NFL) Players Association, for which he receives travel support, and an advisor and options-holder with Oxeia Biopharmaceuticals, LLC, and StataDx; receiving travel support from the NFL, NFL Players Association, World Rugby, World Wrestling Entertainment (WWE), and All Elite Wrestling (AEW); serving as an expert witness in cases related to concussion and chronic traumatic encephalopathy (CTE) and receiving compensation for speaking appearances and serving on the Players Advocacy Committee for the NFL Concussion Settlement; and being employed by the Concussion Legacy Foundation, a 501(c)(3) nonprofit which receives charitable donations.

Dr Tripodis reported receiving personal fees from the American Medical Association outside the submitted work.

Dr Dams-O’Connor reported receiving personal fees from various law firms outside the submitted work.

Dr Katz reported royalties from Springer/Demos Publishers for a textbook on Brain Injury Medicine that includes a chapter on CTE and related disorders.

Dr Stern reported receiving personal fees from Eisai outside the submitted work and royalties for published neuropsychological tests from Psychological Assessment Resources, Inc; owning stock options as a member of Board of Directors for King-Devick Technologies, Inc; receiving grants from the National Institutes of Health (NIH), Life Molecular Imaging Inc, and Rainwater Charitable Foundation Inc during the conduct of the study; and receiving an honorarium from the Michael J. Fox Foundation unrelated to this work and royalties from Oxford University Press Inc outside the submitted work.

Dr Daneshvar reported receiving personal fees for providing expert testimony related to traumatic brain injury and spinal cord injury, serving as a medical advisor and options holder for StataDx, receiving research funding from the Football Players Health Study at Harvard University (FPHS) funded by the NFL Players Association (NFLPA), serving as a volunteer member of the Mackey-White Committee of the NFLPA, and receiving clinical funding from the Brain and Body Program funded by the NFLPA, all outside the submitted work.

## Notes

### Funding Statement

This study was funded by the National Institute of Neurological Disorders and Stroke (U54NS115266, U01NS086659, U01NS093334, R01NS078337, R56NS078337, RF1NS143015, R01NS142076, R01NS122854, RF1NS115268, R01NS128961), National Institute on Aging (P30AG072978, P30AG066514, U19AG068753, R01AG061028, K23AG046377, R21HD089088, F32NS096803, RF1AG062348, R01AG077588), National Heart, Lung and Blood Institute (75N92019D00031), Department of Veterans Affairs (I01 CX001135, CSP 501, B6796-C), Department of Defense (W81XWH-13-2-0095, W81XWH-13-2-0064, W81XWH1810580, PRARP-13267017), the Alzheimers Association (NIRG-15-362697, NIRG-305779), the National Operating Committee on Standards for Athletic Equipment (NOCSAE), the Nick and Lynn Buoniconti Foundation, the Concussion & CTE Foundation, and a gift from William Collatos. The funders had no role in the design and conduct of the study; collection, management, analysis, and interpretation of the data; preparation, review, or approval of the manuscript; and decision to submit the manuscript for publication.

### Author Declarations

The institutional review boards of the Boston University Chobanian and Avedisian School of Medicine, the VA Bedford Healthcare system, the Icahn School of Medicine at Mt. Sinai, and the VA Boston Healthcare System and VA Office of Research and Development gave ethical approval for this work.

## References

1. McKee AC, Stern RA, Nowinski CJ, et al. The spectrum of disease in chronic traumatic encephalopathy. Brain. 2013;136(Pt 1):43–64. doi:10.1093/brain/aws307

2. Mez J, Daneshvar DH, Abdolmohammadi B, et al. Duration of American Football Play and Chronic Traumatic Encephalopathy. Ann Neurol. 2020;87(1):116–131. doi:10.1002/ana.25611

3. Abdolmohammadi B, Tuz-Zahra F, Uretsky M, et al. Duration of Ice Hockey Play and Chronic Traumatic Encephalopathy. JAMA Netw Open. 2024;7(12):e2449106. doi:10.1001/jamanetworkopen.2024.49106

4. Stewart W, Buckland ME, Abdolmohammadi B, et al. Risk of chronic traumatic encephalopathy in rugby union is associated with length of playing career. Acta Neuropathol. 2023;146(6):829–832. doi:10.1007/s00401-023-02644-3

5. Mez J, Daneshvar DH, Kiernan PT, et al. Clinicopathological Evaluation of Chronic Traumatic Encephalopathy in Players of American Football. JAMA. 2017;318(4):360–370. doi:10.1001/jama.2017.8334

6. Goldstein LE, Fisher AM, Tagge CA, et al. Chronic traumatic encephalopathy in blast-exposed military veterans and a blast neurotrauma mouse model. Sci Transl Med. 2012;4(134):134ra60. doi:10.1126/scitranslmed.3003716

7. Alosco ML, Cherry JD, Huber BR, et al. Characterizing tau deposition in chronic traumatic encephalopathy (CTE): utility of the McKee CTE staging scheme. Acta Neuropathol. 2020;140(4):495–512. doi:10.1007/s00401-020-02197-9

8. Ling H, Morris HR, Neal JW, et al. Mixed pathologies including chronic traumatic encephalopathy account for dementia in retired association football (soccer) players. Acta Neuropathol. 2017;133(3):337–352. doi:10.1007/s00401-017-1680-3

9. Bieniek KF, Ross OA, Cormier KA, et al. Chronic traumatic encephalopathy pathology in a neurodegenerative disorders brain bank. Acta Neuropathol. 2015;130(6):877–889. doi:10.1007/s00401-015-1502-4

10. McKee AC, Cairns NJ, Dickson DW, et al. The first NINDS/NIBIB consensus meeting to define neuropathological criteria for the diagnosis of chronic traumatic encephalopathy. Acta Neuropathol. 2016;131(1):75–86. doi:10.1007/s00401-015-1515-z

11. Bieniek KF, Cairns NJ, Crary JF, et al. The Second NINDS/NIBIB Consensus Meeting to Define Neuropathological Criteria for the Diagnosis of Chronic Traumatic Encephalopathy. J Neuropathol Exp Neurol. Published online February 21, 2021. doi:10.1093/jnen/nlab001

12. Stern RA, Daneshvar DH, Baugh CM, et al. Clinical presentation of chronic traumatic encephalopathy. Neurology. 2013;81(13):1122–1129. doi:10.1212/WNL.0b013e3182a55f7f

13. Taghdiri F, Khodadadi M, Sadia N, et al. Unusual combinations of neurodegenerative pathologies with chronic traumatic encephalopathy (CTE) complicates clinical prediction of CTE. European Journal of Neurology. 2024;31(6):e16259. doi:10.1111/ene.16259

14. Asken BM, Tanner JA, VandeVrede L, et al. Multi-Modal Biomarkers of Repetitive Head Impacts and Traumatic Encephalopathy Syndrome: A Clinicopathological Case Series. Journal of Neurotrauma. 2022;39(17-18):1195–1213. doi:10.1089/neu.2022.0060

15. Schaffert J, Iyengar N, Magill R, et al. Clinical and Neuropsychological Profiles in People With Chronic Traumatic Encephalopathy Neuropathologic Change: Matched Case-Series Study. Neurology. 2025;105(11):e214368. doi:10.1212/WNL.0000000000214368

16. Alosco ML, White M, Bell C, et al. Cognitive, functional, and neuropsychiatric correlates of regional tau pathology in autopsy-confirmed chronic traumatic encephalopathy. Mol Neurodegener. 2024;19(1):10. doi:10.1186/s13024-023-00697-2

17. Layden RM, Groh JR, Miner AE, et al. CTE Neuropathology Alone is Associated with Dementia and Cognitive Symptoms. Alzheimer’s & Dementia. Published online 2026. doi:10.1002/alz.71032

18. Jordan B. Medical Aspects of Boxing. Taylor & Francis; 1992. https://books.google.com/books?id=wAlXNUzDUXwC

19. Jordan BD. The clinical spectrum of sport-related traumatic brain injury. Nat Rev Neurol. 2013;9(4):222–230. doi:10.1038/nrneurol.2013.33

20. Victoroff J. Traumatic encephalopathy: review and provisional research diagnostic criteria. NeuroRehabilitation. 2013;32(2):211–224. doi:10.3233/NRE-130839

21. Folstein MF, Nervous A for R in, Disease M. Neurobiology of Primary Dementia. American Psychiatric Press; 1998. https://books.google.com/books?id=TMv5TmYtMlEC

22. Montenigro PH, Baugh CM, Daneshvar DH, et al. Clinical subtypes of chronic traumatic encephalopathy: literature review and proposed research diagnostic criteria for traumatic encephalopathy syndrome. Alzheimer’s Research & Therapy. 2014;6(5):68. doi:10.1186/s13195-014-0068-z

23. Mez J, Alosco ML, Daneshvar DH, et al. Validity of the 2014 traumatic encephalopathy syndrome criteria for CTE pathology. Alzheimer’s & Dementia. 2021;n/a(n/a). doi:10.1002/alz.12338

24. Schaffert J, Didehbani N, LoBue C, et al. Frequency and Predictors of Traumatic Encephalopathy Syndrome in a Prospective Cohort of Retired Professional Athletes. Frontiers in Neurology. 2021;Volume 12-2021. doi:10.3389/fneur.2021.617526

25. Iverson GL, Gardner AJ. Symptoms of traumatic encephalopathy syndrome are common in the US general population. Brain Communications. 2021;3(1):fcab001. doi:10.1093/braincomms/fcab001

26. Katz DI, Bernick C, Dodick DW, et al. National Institute of Neurological Disorders and Stroke Consensus Diagnostic Criteria for Traumatic Encephalopathy Syndrome. Neurology. 2021;96(18):848–863. doi:10.1212/WNL.0000000000011850

27. Mez J, Solomon TM, Daneshvar DH, et al. Assessing clinicopathological correlation in chronic traumatic encephalopathy: rationale and methods for the UNITE study. Alzheimers Res Ther. 2015;7(1):62. doi:10.1186/s13195-015-0148-8

28. Au R, Seshadri S, Knox K, et al. The Framingham Brain Donation Program: neuropathology along the cognitive continuum. Curr Alzheimer Res. 2012;9(6):673–686. doi:10.2174/156720512801322609

29. Alosco ML, Morrison M, Au R, et al. Boston University Alzheimer’s Disease Research Center Clinical Core: Infrastructure to facilitate research on post-traumatic Alzheimer’s disease and related dementias. Alzheimers Dement. 2025;21(9):e70654. doi:10.1002/alz.70654

30. Fischer DL, Grinberg LT, Ahrendsen JT, et al. Celebrating neuropathology’s contributions to Alzheimer’s Disease Research Centers. Alzheimers Dement. 2025;21(10):e70734. doi:10.1002/alz.70734

31. Brady CB, Trevor KT, Stein TD, et al. The Department of Veterans Affairs Biorepository Brain Bank: a national resource for amyotrophic lateral sclerosis research. Amyotroph Lateral Scler Frontotemporal Degener. 2013;14(7-8):591–597. doi:10.3109/21678421.2013.822516

32. Edlow BL, Keene CD, Perl DP, et al. Multimodal Characterization of the Late Effects of Traumatic Brain Injury: A Methodological Overview of the Late Effects of Traumatic Brain Injury Project. J Neurotrauma. 2018;35(14):1604–1619. doi:10.1089/neu.2017.5457

33. Kay T, Harrington D, Adams R, et al. Definition of mild traumatic brain injury. The Journal of Head Trauma Rehabilitation. 1993;8(3):86–87.

34. O’Neil ME, Carlson K, Storzbach D, et al. Complications of Mild Traumatic Brain Injury in Veterans and Military Personnel: A Systematic Review. 2013.

35. Vonsattel JPG, Del Amaya MP, Keller CE. Twenty-first century brain banking. Processing brains for research: the Columbia University methods. Acta Neuropathol. 2008;115(5):509–532. doi:10.1007/s00401-007-0311-9

36. McKee AC, Cantu RC, Nowinski CJ, et al. Chronic traumatic encephalopathy in athletes: progressive tauopathy after repetitive head injury. J Neuropathol Exp Neurol. 2009;68(7):709–735. doi:10.1097/NEN.0b013e3181a9d503

37. Consensus recommendations for the postmortem diagnosis of Alzheimer’s disease. The National Institute on Aging, and Reagan Institute Working Group on Diagnostic Criteria for the Neuropathological Assessment of Alzheimer’s Disease. Neurobiol Aging. 1997;18(4 Suppl):S1–2.

38. McKeith IG. Consensus guidelines for the clinical and pathologic diagnosis of dementia with Lewy bodies (DLB): report of the Consortium on DLB International Workshop. J Alzheimers Dis. 2006;9(3 Suppl):417–423. doi:10.3233/jad-2006-9s347

39. Mackenzie IRA, Neumann M, Bigio EH, et al. Nomenclature for neuropathologic subtypes of frontotemporal lobar degeneration: consensus recommendations. Acta Neuropathol. 2009;117(1):15–18. doi:10.1007/s00401-008-0460-5

40. Nelson PT, Dickson DW, Trojanowski JQ, et al. Limbic-predominant age-related TDP-43 encephalopathy (LATE): consensus working group report. Brain. 2019;142(6):1503–1527. doi:10.1093/brain/awz099

41. Brownell B, Oppenheimer DR, Hughes JT. The central nervous system in motor neurone disease. J Neurol Neurosurg Psychiatry. 1970;33(3):338–357. doi:10.1136/jnnp.33.3.338

42. Corrigan JD, Bogner J. Initial reliability and validity of the Ohio State University TBI Identification Method. J Head Trauma Rehabil. 2007;22(6):318–329. doi:10.1097/01.HTR.0000300227.67748.77

43. Seichepine DR, Stamm JM, Daneshvar DH, et al. Profile of self-reported problems with executive functioning in college and professional football players. J Neurotrauma. 2013;30(14):1299–1304. doi:10.1089/neu.2012.2690

44. Robbins CA, Daneshvar DH, Picano JD, et al. Self-reported concussion history: impact of providing a definition of concussion. Open Access J Sports Med. 2014;5:99–103. doi:10.2147/OAJSM.S58005

45. Bertens LCM, Broekhuizen BDL, Naaktgeboren CA, et al. Use of expert panels to define the reference standard in diagnostic research: a systematic review of published methods and reporting. PLoS Med. 2013;10(10):e1001531. doi:10.1371/journal.pmed.1001531

46. McKee AC, Mez J, Abdolmohammadi B, et al. Neuropathologic and Clinical Findings in Young Contact Sport Athletes Exposed to Repetitive Head Impacts. JAMA Neurol. Published online August 28, 2023. doi:10.1001/jamaneurol.2023.2907

47. Petersen ML, Porter KE, Gruber S, Wang Y, van der Laan MJ. Diagnosing and responding to violations in the positivity assumption. Stat Methods Med Res. 2012;21(1):31–54. doi:10.1177/0962280210386207

48. McGraw KO, Wong SP. Forming inferences about some intraclass correlation coefficients. Psychological methods. 1996;1(1):30.

49. Knopman DS, DeKosky ST, Cummings JL, et al. Practice parameter: diagnosis of dementia (an evidence-based review). Report of the Quality Standards Subcommittee of the American Academy of Neurology. Neurology. 2001;56(9):1143–1153. doi:10.1212/wnl.56.9.1143

50. Rascovsky K, Hodges JR, Knopman D, et al. Sensitivity of revised diagnostic criteria for the behavioural variant of frontotemporal dementia. Brain. 2011;134(Pt 9):2456–2477. doi:10.1093/brain/awr179

51. Harris JM, Thompson JC, Gall C, et al. Do NIA-AA criteria distinguish Alzheimer’s disease from frontotemporal dementia? Alzheimers Dement. 2015;11(2):207–215. doi:10.1016/j.jalz.2014.04.516

52. Ferman TJ, Boeve BF, Smith GE, et al. Inclusion of RBD improves the diagnostic classification of dementia with Lewy bodies. Neurology. 2011;77(9):875–882. doi:10.1212/WNL.0b013e31822c9148

53. Beach TG, Monsell SE, Phillips LE, Kukull W. Accuracy of the clinical diagnosis of Alzheimer disease at National Institute on Aging Alzheimer Disease Centers, 2005-2010. J Neuropathol Exp Neurol. 2012;71(4):266–273. doi:10.1097/NEN.0b013e31824b211b

54. Albert MS, DeKosky ST, Dickson D, et al. The diagnosis of mild cognitive impairment due to Alzheimer’s disease: recommendations from the National Institute on Aging-Alzheimer’s Association workgroups on diagnostic guidelines for Alzheimer’s disease. Alzheimers Dement. 2011;7(3):270–279. doi:10.1016/j.jalz.2011.03.008

55. McKhann GM, Knopman DS, Chertkow H, et al. The diagnosis of dementia due to Alzheimer’s disease: recommendations from the National Institute on Aging-Alzheimer’s Association workgroups on diagnostic guidelines for Alzheimer’s disease. Alzheimers Dement. 2011;7(3):263–269. doi:10.1016/j.jalz.2011.03.005

56. McKeith IG, Boeve BF, Dickson DW, et al. Diagnosis and management of dementia with Lewy bodies: Fourth consensus report of the DLB Consortium. Neurology. 2017;89(1):88–100. doi:10.1212/WNL.0000000000004058

57. Knopman DS, Petersen RC. Mild cognitive impairment and mild dementia: a clinical perspective. Mayo Clin Proc. 2014;89(10):1452–1459. doi:10.1016/j.mayocp.2014.06.019

58. Sperling RA, Aisen PS, Beckett LA, et al. Toward defining the preclinical stages of Alzheimer’s disease: recommendations from the National Institute on Aging-Alzheimer’s Association workgroups on diagnostic guidelines for Alzheimer’s disease. Alzheimers Dement. 2011;7(3):280–292. doi:10.1016/j.jalz.2011.03.003

59. Frigerio R, Fujishiro H, Ahn TB, et al. Incidental Lewy body disease: do some cases represent a preclinical stage of dementia with Lewy bodies? Neurobiol Aging. 2011;32(5):857–863. doi:10.1016/j.neurobiolaging.2009.05.019

60. Arena JD, Stewart W, Schneider ALC, et al. Performance of traumatic encephalopathy syndrome criteria in identifying individuals with chronic traumatic encephalopathy. Nature Medicine. Published online May 14, 2026. doi:10.1038/s41591-026-04392-9

61. Deeks JJ, Altman DG. Diagnostic tests 4: likelihood ratios. BMJ. 2004;329(7458):168–169. doi:10.1136/bmj.329.7458.168

62. Dams-O’Connor K, Seifert AC, Crary JF, et al. The neuropathology of intimate partner violence. Acta Neuropathol. 2023;146(6):803–815. doi:10.1007/s00401-023-02646-1

