## Supplement packet for "Validity of the NINDS traumatic encephalopathy syndrome criteria for predicting chronic traumatic encephalopathy"

### Supplemental Online Content

#### Table of Contents


| | |
|---|---|
| **Page 2-4** | **eMethods** |
| **Page 5** | **eTable 1. Questionnaires administered across brain banks** |
| **Page 6-10** | **eTable 2. Descriptive statistics and neuropathology by brain bank** |
| **Page 11-16** | **eTable 3. Descriptive statistics and neuropathology by exposure bins** |
| **Page 17-21** | **eTable 4. Descriptive statistics and neuropathology by age bins** |
| **Page 22-23** | **eTable 5. TES consensus diagnosis by CTE pathology sensitivity analyses** |
| **Page 24-25** | **eTable 6. TES criteria, stratified by CTE Stage** |
| **Page 26** | **eFigure 1. Standards for the Reporting of Diagnostic Accuracy Studies (STARD) flow diagram** |
| **Page 27-28** | **eFigure 2. Consensus checklist clinical diagnoses beyond TES** |
| **Page 29** | **References** |

**eMethods**

**Brain Banks**

***UNITE Brain Bank***

For the current study, recruitment from the UNITE Brain Bank occurred between 2021-2024. Methods for data collection and neuropathological assessment have been described previously.[1] To be eligible for donation, donors needed to have a history of RHI exposure from CCS, military service with or without combat exposure, physical violence (i.e. intimate partner violence) or moderate to severe TBI. Potential donors were excluded if the post-mortem interval exceeded 72 hours, or if insufficient tissue was available (i.e., less than one intact hemisphere). Donors' next-of-kin provided written consent for donation and all relevant research activities, including permission to publish deidentified results. Institutional review board (IRB) approval was obtained through the Boston University Chobanian and Avedisian School of Medicine and the VA Bedford Healthcare system.

***Framingham Heart Study***

The FHS was established in 1948, enrolling a representative group of residents from Framingham, Massachusetts. Over time, the study expanded to include their children (Gen 2) and grandchildren (Gen 3). Additionally, ethnically diverse cohorts (Omni 1, Omni 2) were added to reflect the changing demographics of Framingham. The FHS Brain Bank, a voluntary subgroup of participants, began enrolling members in 1997.[2] Criteria for inclusion into the FHS Brain Bank have been previously described.[2] Potential donors were excluded if the post-mortem interval exceeded 72 hours, or if insufficient tissue was available (i.e., less than one intact hemisphere). Donors' next-of-kin provided written consent for donation and all relevant research activities, including permission to publish deidentified results. Institutional review board (IRB) approval was obtained through the Boston University Chobanian and Avedisian School of Medicine.

***BU ADRC***

The BU ADRC was established through funding by the NIA in 1996 and has sustained a mission to facilitate leading-edge research on AD/ADRD. All brain donors were followed in life as part of the BU ADRC Clinical Core. When the BU ADRC was founded, the Center focused on late-stage dementia. As AD/ADRD research evolved, the Center's focus shifted to early-stage disease detection, resulting in a longitudinal cohort of participants who span the continuum of AD (normal cognition [NC], mild cognitive impairment [MCI], and dementia). Since 2018, the Center began to integrate examination of participants exposed to RHI with emphasis on the comparison of those with and without RHI. Potential donors were excluded if the post-mortem interval exceeded 72 hours, or if insufficient tissue was available (i.e., less than one intact hemisphere). Donors' next-of-kin provided written consent for donation and all relevant research activities, including permission to publish deidentified results. Institutional review board (IRB) approval was obtained through the Boston University Chobanian and Avedisian School of Medicine[3].

***Mt. Sinai ADRC***

The Alzheimer's Disease Research Center at the Icahn School of Medicine at Mt. Sinai was established in 1984 to contribute to knowledge about AD and other dementias. All brain donors were followed in life as part of the Mt. Sinai ADRC Clinical Core. Willing participants were enrolled in the Brain Tissue Donation Program; upon death, ADRC staff recovered the brain tissue from decedents. A comprehensive neuropathology report was prepared, detailing diagnoses and findings from donated brain tissue, typically completed 6-12 months following death. Donors' next-of-kin provided written consent for donation and all relevant research activities, including permission to publish deidentified results. Institutional review board (IRB) approval was obtained through the Icahn School of Medicine[4].

***LETBI***
The Late Effects of Traumatic Brain Injury (LETBI) study was established in 2014 to examine clinical, imaging, and histopathological signatures of neurodegenerative disease in individuals with a history of traumatic brain injury (TBI). Donors' next-of-kin provided written consent for donation and all relevant research activities, including permission to publish deidentified results. Institutional review board (IRB) approval was obtained through the Icahn School of Medicine[5].

***VA ALS***
The Veterans Affairs Biorepository Brain Bank (VABBB) was established in 2006. The VABBB is a human tissue bank collecting and storing brain and spinal cord tissue, with the goal of encouraging research into amyotrophic lateral sclerosis (ALS), primary lateral sclerosis, progressive bulbar palsy, and progressive muscular atrophy in former military veterans. Veterans in the United States with ALS, ALS-related conditions, or those without neurological disease in the U.S. were eligible for enrollment in the VABBB. A subset of brain donors were followed during life. Legal next-of-kin were asked to sign provisional consent forms agreeing to make an after-death organ donation. Institutional review board (IRB) approval was obtained through the VA Boston Healthcare System and VA Office of Research and Development[6].

**Traumatic encephalopathy syndrome diagnostic criteria**
TES was diagnosed utilizing the most recent iteration of the TES diagnostic criteria as published by Katz et al. in 2021.[7] Four primary criteria are necessary for TES diagnosis.

I) Substantial exposure to repetitive head impacts: a history of high-level contact sport play (i.e., 5+ years of American football play or equivalent), military service with a history of RHI, intimate partner violence or childhood physical violence. Donors with substantial exposure were further classified as having had either extensive or less than extensive exposure, based on pre-defined exposure risk thresholds.[7–9]
II) Core clinical features
  a. Cognitive impairment: presence of deficits in episodic memory and/or executive function, representing a significant decline from baseline.
  b. Neurobehavioral dysregulation: presence of symptoms or observed behaviors representing poor regulation of emotions and/or behavior, including emotional lability, explosiveness, impulsivity, short fuse, or violent outbursts. These symptoms should not be transient responses to life events (i.e. divorce, death of loved ones). Symptoms should represent a significant decline from baseline.
  c. Progressive course: Evidence of progressive worsening of one or both of the clinical features over a period of at minimum one year, in the absence of exposure to RHI. If available, evidence should be supported by standardized neuropsychological testing and/or physician records or self or informant-reports.
III) Not fully accounted for by other disorders: A pattern of cognitive deficits which are not fully accounted for by other preexisting or established nondegenerative nervous system, psychiatric, substance use, or medical disorders or conditions.
IV) Level of functional dependence: A qualitative assessment based on the impact of cognitive impairment and/or neurobehavioral dysregulation. Ranges from fully independent to severe dementia. Level of functional dependence must represent a change from previous baseline functioning

Additional supportive features include:

- Delayed onset of symptoms: Core clinical features begin following a clearly established period of stable functioning after the RHI exposure ends. (A minimum time period of stability before onset and progression of symptoms has not been established but should be substantial [i.e., years] to suggest a history consistent with a degenerative disorder rather than problems associated with TBI or other preexisting conditions.)

- Motor signs
  - Parkinsonism: bradykinesia, rigidity, rest tremor, and parkinsonian gait disorder; these motor signs should not be more consistent with the clinical features of nonparkinsonian neurologic conditions or primary orthopedic problems.
  - Other motor signs: dysarthria, ataxia, and imbalance; these motor signs should not be more consistent with the clinical features of other neurologic conditions or primary orthopedic problems.
  - Motor neuron disease: Weakness, dysphagia, other lower motor neuron signs (fasciculations and muscle atrophy), and other upper motor neuron signs (spasticity, hyperreflexia, extensor plantar response, and spastic dysarthria); a diagnosis of amyotrophic lateral sclerosis (ALS) would meet this criterion but is not necessary.
- Presence of psychiatric features: These supportive psychiatric features have not been accounted for by neurobehavioral dysregulation described in the core clinical features. They may occur individually or in combination, should represent a clear change from baseline, and should be persistent (i.e., months to years) or progressive. These features can be based on self- or informant report, a history of treatment, or clinician's report. The supportive psychiatric features include the following
  - Anxiety: Pervasive worries, excessive fears, agitation, or obsessive or compulsive behavior (or both); a formal diagnosis of anxiety disorder would meet this criterion but is not necessary. If available, scores on an established, validated anxiety scale should indicate a moderate level of anxiety or higher.
  - Apathy: Loss of interest in usual activities and loss of motivation or drive. If available, scores on an established, validated apathy scale should indicate a moderate level of apathy or higher.
  - Depression: Feeling overly sad, dysphoric, or hopeless, with or without a history of suicidal thoughts or attempts; a formal diagnosis of major depressive disorder or persistent depressive disorder would meet this criterion but is not necessary. These symptoms should not be a time-limited reaction to an event (e.g., death of family member, illness, and trauma). If available, scores on an established, validated depression scale should indicate a moderate level of depression or higher.
  - Paranoia: Delusional beliefs of suspicion, persecution, or unwarranted jealousy; a formal diagnosis of a psychotic disorder would meet this criterion but is not necessary. If available, scores on an established, validated paranoia scale should indicate a moderate level of paranoia or higher.[7]

**eTable 1. Questionnaires administered across brain banks**

| Brain Bank | Data collected |
|---|---|
| UNITE | Athletic and military histories (taken from BU Head Impact Exposure Assessment); Clinical Scales (FAQ, GDS, CDS, BRIEF-A, AES, BIS-11, BAI, BGS, NPIQ); Ohio State University TBI survey |
| BU ADRC | Athletic and military histories (taken from BU Head Impact Exposure Assessment); Clinical Scales (FAQ, GDS, CDS, BRIEF-A, AES, BIS-11, BAI, BGS, NPIQ); Ohio State University TBI survey |
| FHS | Athletic and military histories (taken from BU Head Impact Exposure Assessment); Clinical Scales (FAQ, GDS, CDS, NPIQ); Ohio State University TBI survey |
| Mt. Sinai ADRC | Athletic and military histories (taken from BU Head Impact Exposure Assessment); Clinical Scales (FAQ, BGS, NPIQ); Brain Injury Screening Questionnaire (BISQ) |
| LETBI | Athletic and military histories (taken from BU Head Impact Exposure Assessment); Clinical Scales (FAQ, BGS, NPIQ); Brain Injury Screening Questionnaire (BISQ) |
| VA ALS | Athletic and military histories (taken from BU Head Impact Exposure Assessment); Clinical Scales (FAQ, CDS, NPIQ); Ohio State University TBI survey |

**eTable 2. Descriptives and Neuropathology (stratified by brain bank)**

| | UNITE | FHS | BU ADRC | Mt.Sinai ADRC | VA ALS | LETBI |
|---|---|---|---|---|---|---|
| **Number (%)** | 86 (44.3) | 32 (17) | 25 (12.9) | 16 (8.2) | 16 (8.2) | 18 (9.3) |
| **Demographics** | | | | | | |
| Age at death (years) | 51.31 ± 20.35 | 83.55 ± 12.58 | 83.92 ± 10.54 | 82.5 ± 11.81 | 72.81 ± 9.83 | 63.72 ± 17.31 |
| Female sex | 3 (3.5) | 11 (33.3) | 12 (48) | 11 (68.8) | 1 (6.3) | 2 (11.1) |
| Race | | | | | | |
| Black | 10 (11.6) | 0 | 1 (4) | 2 (12.5) | 0 | 2 (11.1) |
| White | 70 (81.4) | 32 (97) | 23 (92) | 8 (50) | 14 (87.5) | 12 (66.7) |
| Multiple | 5 (5.8) | 0 | 0 | 0 | 0 | 0 |
| Other | 1 (1.2) | 0 | 0 | 0 | 0 | 0 |
| Unreported | 0 | 1 (3) | 1 (4) | 6 (37.5) | 2 (12.5) | 4 (22.2) |
| **Donor Recruitment categories** | | | | | | |
| RHI exposure | | | | | | |
| College Football< | 12 (14) | 6 (18.2) | 2 (8) | 1 (6.3) | 2 (12.5) | 2 (11.1) |
| College Football+ | 33 (38.4) | 0 | 2 (8) | 0 | 1 (6.3) | 1 (5.6) |
| College contact sport< | 9 (10.4) | 3 (9.1) | 2 (8) | 0 | 1 (6.3) | 2 (11.1) |
| College contact sport+ | 18 (20.9) | 2 (6.1) | 0 | 0 | 0 | 1 (5.6) |
| Military w/combat, no sport | 2 (2.3) | 7 (21.2) | 5 (20) | 1 (6.3) | 4 (25) | 0 |
| Military w/combat + sport | 9 (10.5) | 0 | 0 | 0 | 4 (25) | 2 (11.1) |
| 1+ mTBI w/LOC, no sport/military | 1 (1.2) | 6 (18.2) | 3 (12) | 0 | 3 (18.8) | 2 (11.1) |
| 1+ mod-severe TBI, no sport/military | 2 (2.3) | 4 (12.1) | 0 | 1 (6.3) | 0 | 8 (44.4) |
| No RHI exposure | 0 | 5 (15.2) | 11 (44) | 13 (81.3) | 1 (6.3) | 0 |
| **Age** | | | | | | |
| 20-34 | 24 (27.9) | 0 | 0 | 0 | 0 | 1 (5.6) |
| 35-49 | 22 (25.6) | 0 | 0 | 0 | 0 | 3 (16.7) |
| 50-64 | 16 (18.6) | 5 (15.6) | 1 (4) | 0 | 4 (25) | 3 (16.7) |
| 65-79 | 11 (12.8) | 3 (9.4) | 6 (24) | 7 (43.8) | 8 (50) | 8 (44.4) |
| 80+ | 13 (15.1) | 24 (75) | 18 (72) | 9 (56.3) | 4 (25) | 3 (16.7) |

| | UNITE | FHS | BU ADRC | Mt.Sinai ADRC | VA ALS | LETBI |
|---|---|---|---|---|---|---|
| **TES Criteria** | | | | | | |
| **TES status** | | | | | | |
| No | 24 (27.9) | 30 (90.9) | 23 (92) | 16 (100) | 12 (75) | 16 (88.9) |
| Suggestive | 12 (14) | 1 (3) | 0 | 0 | 1 (6.3) | 1 (5.6) |
| Possible | 21 (24.4) | 1 (3) | 2 (8) | 0 | 3 (18.8) | 1 (5.6) |
| Probable | 29 (33.7) | 0 | 0 | 0 | 0 | 0 |
| **Substantial exposure to RHI** | 83 (96.5) | 17 (53.1) | 5 (20) | 0 | 12 (75) | 12 (66.7) |
| **Extensive exposure to RHI** | 37 (43) | 0 | 0 | 0 | 0 | 0 |
| **Cognitive impairment** | 67 (77.9) | 18 (54.5) | 22 (88) | 15 (93.8) | 9 (56.3) | 12 (66.7) |
| **Neurobehavioral dysregulation** | 78 (90.7) | 6 (18.2) | 6 (24) | 7 (43.8) | 10 (62.5) | 9 (50) |
| **Progressive course** | | | | | | |
| Yes | 78 (90.7) | 19 (57.6) | 22 (88) | 15 (93.8) | 11 (68.8) | 13 (72.2) |
| No | 1 (1.2) | 0 | 2 (8) | 0 | 0 | 0 |
| N/A | 1 (1.2) | 13 (40.6) | | 1 (6.3) | 5 (31.3) | 4 (22.2) |
| Inconclusive | 6 (7) | 0 | 1 (4) | 0 | 0 | 1 (5.6) |
| **Fully Accounted for by Other Disorders** | | | | | | |
| No | 67 (77.9) | 3 (9.4) | 4 (16) | 0 | 4 (25) | 2 (11.1) |
| Yes | 15 (17.4) | 14 (43.8) | 9 (36) | 15 (93.8) | 7 (43.8) | 12 (66.7) |
| N/A | 4 (4.7) | 15 (46.9) | 12 (48) | 1 (6.3) | 5 (31.3) | 4 (22.2) |
| **Level of Functional Dependence/Dementia** | | | | | | |
| Independent/no dementia | 11 (12.8) | 15 (45.5) | 3 (12) | 1 (6.3) | 5 (31.3) | 5 (27.8) |
| Subtle functional limitation | 36 (41.9) | 3 (9.1) | 1 (4) | 0 | 5 (31.3) | 2 (11.1) |
| Mild dementia | 9 (10.5) | 2 (6.1) | 3 (12) | 1 (6.3) | 3 (18.8) | 4 (22.2) |
| Moderate dementia | 10 (11.6) | 2 (6.1) | 5 (20) | 1 (6.3) | 2 (12.5) | 3 (16.7) |
| Severe dementia | 20 (23.3) | 10 (30.3) | 13 (52) | 13 (81.3) | 1 (6.3) | 4 (22.2) |
| **Delayed onset (of one or more core clinical features)** | 56 (65.1) | 3 (9.1) | 4 (16) | 0 | 7 (43.8) | 5 (27.8) |
| **Parkinsonism** | 22 (25.6) | 8 (24.2) | 8 (32) | 3 (18.8) | 2 (12.5) | 5 (27.8) |
| **Other Motor Signs** | 7 (8.1) | 0 | 1 (4) | 1 (6.3) | 0 | 2 (11.1) |
| **Motor Neuron Disease** | 1 (1.2) | 1 (3) | 0 | 0 | 15 (93.8) | 0 |

| | UNITE | FHS | BU ADRC | Mt.Sinai ADRC | VA ALS | LETBI |
|---|---|---|---|---|---|---|
| **Psychiatric Features** | 85 (98.8) | 16 (48.5) | 21 (84) | 12 (75) | 12 (75) | 15 (83.3) |
| Anxiety | 60 (69.8) | 7 (21.2) | 9 (36) | 3 (18.8) | 3 (18.8) | 10 (55.6) |
| Apathy | 73 (84.9) | 5 (15.2) | 18 (72) | 10 (62.5) | 6 (37.5) | 10 (55.6) |
| Depression | 70 (81.4) | 12 (36.4) | 10 (40) | 7 (43.8) | 8 (50) | 11 (61.1) |
| Paranoia | 53 (61.6) | 5 (15.2) | 4 (16) | 3 (18.8) | 4 (25) | 7 (38.9) |
| **Additional clinical information** | | | | | | |
| Neuropsychological testing | 47 (54.7) | 32 (100) | 24 (96) | 11 (68.8) | 0 | 9 (50) |
| MRI | 14 (16.3) | 19 (59.4) | 2 (8) | 11 (68.8) | 1 (6.3) | 5 (27.8) |
| AD biomarkers (either CSF or amyloid PET) | 5 (5.8) | 2 (6.3) | 1 (4) | 0 | 0 | 0 |
| **Neuropathology** | | | | | | |
| **CTE** | 49 (57) | 1 (3.1) | 0 | 0 | 5 (31.3) | 2 (11.1) |
| **CTE Stage** | | | | | | |
| Stage I | 9 (10.5) | 1 (3.1) | 0 | 0 | 3 (18.8) | 1 (5.6) |
| Stage II | 10 (11.6) | 0 | 0 | 0 | 1 (6.3) | 0 |
| Stage III | 14 (16.3) | 0 | 0 | 0 | 1 (6.3) | 1 (5.6) |
| Stage IV | 16 (18.6) | 0 | 0 | 0 | 0 | 0 |
| **CTE II-IV** | 40 (46.5) | 0 | 0 | 0 | 2 (12.5) | 1 (5.6) |
| **Alzheimer's disease NIA-Reagan criteria** | 22 (25.6) | 13 (40.6) | 13 (52) | 12 (75) | 3 (18.8) | 6 (33.3) |
| **FTLD diagnosis** | 3 (3.5) | 5 (15.6) | 3 (12) | 0 | 5 (31.3) | 1 (5.6) |
| **Lewy body disease** | | | | | | |
| Brainstem predominant | 2 (2.3) | 4 (12.5) | 3 (12) | 4 (25) | 0 | 4 (22.2) |
| Limbic (transitional) | 7 (8.1) | 2 (6.3) | 2 (8) | 0 | 0 | 0 |
| Neocortical (diffuse) | 0 | 0 | 2 (8) | 0 | 0 | 1 (5.6) |
| Olfactory bulb | 0 | 0 | 1 (4) | 0 | 0 | 0 |
| **Motor neuron disease** | | | | | | |
| With TDP-43 inclusions in motor neurons | 1 (1.2) | 0 | 0 | 0 | 15 (93.8) | 0 |
| With other inclusions | 0 | 1 (3.1) | 0 | 0 | 0 | 0 |
| **Thal phase (n=163)** | | | | | | |
| Phase 1 (A1) | 2 (2.3) | 1 (6.7) | 2 (16.7) | 1 (6.3) | 7 (43.8) | 2 (11.1) |
| Phase 2 (A1) | 1 (1.2) | 0 | 2 (16.7) | 2 (12.5) | 0 | 0 |
| Phase 3 (A2) | 9 (10.5) | 5 (33.3) | 1 (8.3) | 2 (12.5) | 2 (12.5) | 4 (22.2) |
| Phase 4 (A3) | 11 (12.8) | 4 (26.7) | 3 (25) | 5 (31.3) | 1 (6.3) | 4 (22.2) |
| Phase 5 (A3) | 10 (11.6) | 2 (13.3) | 4 (33.3) | 4 (25) | 1 (6.3) | 1 (5.6) |

| | UNITE | FHS | BU ADRC | Mt.Sinai ADRC | VA ALS | LETBI |
|---|---|---|---|---|---|---|
| **Braak stage (n=180)** | | | | | | |
| Stage I (B1) | 13 (15.1) | 5 (15.6) | 0 | 0 | 2 (12.5) | 2 (11.1) |
| Stage II (B1) | 1 (1.2) | 4 (12.5) | 3 (25) | 2 (12.5) | 4 (25) | 2 (11.1) |
| Stage III (B2) | 6 (7) | 10 (31.3) | 2 (16.7) | 1 (6.3) | 3 (18.8) | 2 (11.1) |
| Stage IV (B2) | 6 (7) | 4 (12.5) | 1 (8.3) | 0 | 2 (12.5) | 4 (22.2) |
| Stage V (B3) | 12 (14) | 1 (3.1) | 4 (33.3) | 7 (43.8) | 0 | 2 (11.1) |
| Stage VI (B3) | 8 (9.3) | 5 (15.6) | 2 (16.7) | 5 (31.3) | 1 (6.3) | 2 (11.1) |
| **CERAD score (neuritic plaques) (n=180)** | | | | | | |
| Sparse neuritic plaques (C1) | 8 (9.3) | 8 (25) | 6 (50) | 1 (6.3) | 5 (31.3) | 5 (27.8) |
| Moderate neuritic plaques (C2) | 11 (12.8) | 8 (25) | 2 (16.7) | 5 (31.3) | 0 | 4 (22.2) |
| Frequent neuritic plaques (C3) | 6 (7) | 1 (3.1) | 2 (16.7) | 8 (50) | 1 (6.3) | 1 (5.6) |
| **CERAD score (diffuse plaques) (n=180)** | | | | | | |
| Sparse diffuse plaques | 5 (5.8) | 5 (15.6) | 4 (33.3) | 1 (6.3) | 6 (37.5) | 5 (27.8) |
| Moderate diffuse plaques | 12 (14) | 6 (18.8) | 4 (33.3) | 5 (31.3) | 3 (18.8) | 4 (22.2) |
| Frequent diffuse plaques | 16 (18.6) | 14 (43.8) | 4 (33.3) | 8 (50) | 2 (12.5) | 1 (5.6) |
| **Arteriolosclerosis (n=180)** | | | | | | |
| Mild | 31 (36) | 4 (12.5) | 1 (8.3) | 13 (81.3) | 8 (50) | 7 (38.9) |
| Moderate | 24 (27.9) | 17 (53.1) | 7 (58.3) | 1 (6.3) | 6 (37.5) | 3 (16.7) |
| Severe | 9 (10.5) | 8 (25) | 3 (25) | 1 (6.3) | 1 (6.3) | 3 (16.7) |
| **Atherosclerosis (n=177)** | | | | | | |
| Mild | 15 (17.4) | 14 (43.8) | 5 (41.7) | 6 (37.5) | 6 (37.5) | 8 (44.4) |
| Moderate | 7 (8.1) | 6 (18.8) | 1 (8.3) | 1 (6.3) | 2 (12.5) | 4 (22.2) |
| Severe | 2 (2.3) | 5 (15.6) | 2 (16.7) | 2 (12.5) | 0 | 0 |
| **White Matter Rarefaction (n=146)** | | | | | | |
| Mild | 32 (37.2) | 13 (40.6) | 5 (41.7) | ~ | 10 (62.5) | ~ |
| Moderate | 33 (38.4) | 4 (12.5) | 3 (25) | ~ | 5 (31.3) | ~ |
| Severe | 3 (3.5) | 4 (12.5) | 2 (16.7) | ~ | 0 | ~ |
| **Cerebral Amyloid Angiopathy (CAA) (n=180)** | | | | | | |
| Mild | 7 (8.1) | 10 (31.3) | 3 (25) | 3 (18.8) | 3 (18.8) | 2 (11.1) |
| Moderate | 8 (9.3) | 10 (33.3) | 1 (8.3) | 5 (31.3) | 2 (12.5) | 0 |
| Severe | 3 (3.5) | 3 (9.4) | 4 (33.3) | 2 (12.5) | 1 (6.3) | 3 (16.7) |
| **Old microinfarcts (n=180)** | 10 (11.6) | 13 (40.6) | 4 (33.3) | 2 (12.5) | 5 (31.3) | 5 (27.8) |
| **Old cerebral microbleeds (n=163)** | 1 (1.2) | 0 | 2 (16.7) | 2 (12.5) | 0 | 8 (44.4) |

| | UNITE | FHS | BU ADRC | Mt.Sinai ADRC | VA ALS | LETBI |
|---|---|---|---|---|---|---|
| **TDP-43 inclusions (n=181)** | 18 (20.9) | 2 (6.3) | 5 (20) | 5 (31.3) | 14 (87.5) | 3 (16.7) |
| Amygdala | 16 (18.6) | 1 (3.1) | 5 (20) | 4 (25) | 7 (43.8) | 1 (5.6) |
| Hippocampus | 16 (18.6) | 1 (3.1) | 4 (16) | 1 (12.5) | 8 (50) | 2 (11.1) |
| Neocortex | 11 (12.8) | 0 | 0 | 0 | 11 (68.8) | 2 (11.1) |

**eTable 3. Descriptives and Neuropathology (stratified by exposure bins)**

| | College FB< | College FB+ | College sport< | College Sport+ | Military w/combat, no sport | Military w/combat+ sport | 1+mTBI w/LOC | 1+ mod-severe TBI | No RHI exposure |
|---|---|---|---|---|---|---|---|---|---|
| **Number (%)** | 25 | 37 | 17 | 21 | 19 | 15 | 15 | 15 | 29 |
| **Demographics** | | | | | | | | | |
| Age at death (years) | 58.24 ± 23.99 | 60.73 ± 21.05 | 54.71 ± 28.05 | 58.33 ± 16.91 | 77.21 ± 15.08 | 51.93 ± 17.98 | 81.93 ± 14.71 | 71.07 ± 21.22 | 83.37 ± 10.07 |
| Female sex | 0 | 0 | 2 (11.8) | 1 (4.8) | 0 | 1 (6.7) | 9 (60) | 5 (33.3) | 22 (73.3) |
| Race | | | | | | | | | |
| Black | 2 (8) | 7 (18.9) | 0 | 1 (4.8) | 0 | 0 | 0 | 2 (13.3) | 3 (10) |
| White | 22 (88) | 27 (73) | 15 (88.2) | 20 (95.2) | 15 (78.9) | 13 (86.7) | 15 (100) | 11 (73.3) | 21 (70) |
| Multiple | 0 | 2 (5.4) | 1 (5.9) | 0 | 1 (5.3) | 1 (6.7) | 0 | 0 | 0 |
| Other | 0 | 0 | 1 (5.9) | 0 | 0 | 0 | 0 | 0 | 0 |
| Unreported | 1 (4) | 1 (2.7) | 0 | 0 | 3 (15.8) | 1 (6.7) | 0 | 2 (13.3) | 6 (20) |
| **Donor Recruitment categories** | | | | | | | | | |
| Brain Bank | | | | | | | | | |
| UNITE | 12 (48) | 33 (89.2) | 9 (52.9) | 18 (85.7) | 2 (10.5) | 9 (60) | 1 (6.7) | 2 (13.3) | 0 |
| FHS | 6 (24) | 0 | 3 (17.6) | 2 (9.5) | 7 (36.8) | 0 | 6 (40) | 4 (26.7) | 5 (16.7) |
| BU ADRC | 2 (8) | 2 (5.4) | 2 (11.8) | 0 | 5 (26.3) | 0 | 3 (20) | 0 | 11 (36.7) |
| Mt. Sinai ADRC | 1 (4) | 0 | 0 | 0 | 1 (5.3) | 0 | 0 | 1 (6.7) | 13 (43.3) |
| VA ALS | 2 (8) | 1 (2.7) | 1 (5.9) | 0 | 4 (21.1) | 4 (26.7) | 3 (20) | 0 | 1 (3.3) |
| LETBI | 2 (8) | 1 (2.7) | 2 (11.8) | 1 (4.8) | 0 | 2 (13.3) | 2 (13.3) | 8 (53.3) | 0 |
| Age | | | | | | | | | |
| 20-34 | 7 (28) | 8 (21.6) | 4 (23.5) | 2 (9.5) | 0 | 2 (13.3) | 0 | 2 (13.3) | 0 |
| 35-49 | 3 (12) | 3 (8.1) | 6 (35.3) | 5 (23.8) | 2 (10.5) | 6 (40) | 0 | 0 | 0 |
| 50-64 | 4 (16) | 8 (21.6) | 0 | 8 (38.1) | 1 (5.3) | 2 (13.3) | 3 (20) | 3 (20) | 0 |
| 65-79 | 6 (24) | 8 (21.6) | 2 (11.8) | 1 (4.8) | 5 (26.3) | 4 (26.7) | 3 (20) | 3 (20) | 11 (37.9) |
| 80+ | 5 (20) | 10 (27) | 5 (29.4) | 5 (23.8) | 11 (57.9) | 1 (6.7) | 9 (60) | 7 (46.7) | 18 (62.1) |

| | College FB< | College FB+ | College sport< | College Sport+ | Military w/combat, no sport | Military w/combat+ sport | 1+mTBI w/LOC | 1+ mod-severe TBI | No RHI exposure |
|---|---|---|---|---|---|---|---|---|---|
| **TES Criteria** | | | | | | | | | |
| **TES status** | | | | | | | | | |
| No | 17 (68) | 7 (18.9) | 12 (70.6) | 5 (23.8) | 17 (89.5) | 4 (26.7) | 15 (100) | 15 (100) | 30 (100) |
| Suggestive | 3 (12) | 2 (5.4) | 2 (11.8) | 1 (4.8) | 2 (10.5) | 5 (33.3) | 0 | 0 | 0 |
| Possible | 5 (20) | 6 (16.2) | 3 (17.6) | 8 (38.1) | 0 | 6 (40) | 0 | 0 | 0 |
| Probable | 0 | 22 (59.5) | 0 | 7 (33.3) | 0 | 0 | 0 | 0 | 0 |
| **Substantial exposure to RHI** | 21 (84) | 37 (100) | 16 (94.1) | 20 (95.2) | 9 (47.4) | 15 (100) | 5 (33.3) | 4 (26.7) | 2 (6.9) |
| **Extensive exposure to RHI** | 0 | 29 (78.4) | 0 | 7 (33.3) | 0 | 1 (6.7) | 0 | 0 | 0 |
| **Cognitive impairment** | 13 (52) | 30 (81.1) | 14 (82.4) | 17 (81) | 13 (68.4) | 11 (73.3) | 12 (80) | 10 (66.7) | 23 (76.7) |
| **Neurobehavioral dysregulation** | 14 (56) | 32 (86.5) | 13 (76.5) | 17 (81) | 8 (42.1) | 13 (86.7) | 5 (33.3) | 4 (26.7) | 10 (33.3) |
| **Progressive course** | | | | | | | | | |
| Yes | 18 (72) | 33 (89.2) | 14 (82.4) | 19 (90.5) | 16 (84.2) | 13 (86.7) | 12 (80) | 10 (66.7) | 23 (76.7) |
| No | 0 | 1 (2.7) | 0 | 0 | 0 | 0 | 0 | 0 | 0 |
| N/A | 7 (28) | 1 (2.7) | 1 (5.9) | 1 (4.8) | 3 (15.8) | 0 | 3 (20) | 5 (33.3) | 5 (17.2) |
| Inconclusive | 0 | 2 (5.4) | 2 (11.8) | 1 (4.8) | 0 | 2 (13.3) | 0 | 0 | 1 (3.3) |
| **Fully Accounted for by Other Disorders** | | | | | | | | | |
| No | 9 (36) | 32 (86.5) | 6 (35.3) | 17 (81) | 3 (15.8) | 12 (80) | 1 (6.7) | 0 | 0 |
| Yes | 8 (32) | 4 (10.8) | 9 (52.9) | 2 (9.5) | 10 (52.6) | 3 (20) | 11 (73.3) | 9 (60) | 16 (55.2) |
| N/A | 8 (32) | 1 (2.7) | 2 (11.8) | 2 (9.5) | 6 (31.6) | 0 | 3 (20) | 6 (40) | 13 (44.8) |
| **Level of Functional Dependence/Dementia** | | | | | | | | | |
| Independent/no | | | | | | 3 (20) | 3 (20) | 4 (26.7) | 6 (20) |
| dementia | 10 (40) | 4 (10.8) | 3 (17.6) | 3 (14.3) | 4 (21.1) | 7 (46.7) | 3 (20) | 2 (13.3) | 1 (3.3) |
| Subtle functional | 7 (28) | 9 (24.3) | 7 (41.2) | 5 (23.8) | 6 (31.6) | 2 (13.3) | 3 (20) | 1 (6.7) | 2 (6.7) |
| limitation | 2 (8) | 5 (13.5) | 1 (5.9) | 4 (19) | 2 (10.5) | 1 (6.7) | 2 (13.3) | 4 (26.7) | 3 (10) |
| Mild dementia | 1 (4) | 6 (16.2) | 3 (17.6) | 2 (9.5) | 1 (5.3) | 2 (13.3) | 4 (26.7) | 4 (26.7) | 17 (56.7) |
| Moderate dementia | 5 (20) | 13 (35.1) | 3 (17.6) | 7 (33.3) | 6 (31.6) | | | | |
| Severe dementia | | | | | | | | | |

| | College FB< | College FB+ | College sport< | College Sport+ | Military w/combat, no sport | Military w/combat+ sport | 1+mTBI w/LOC | 1+ mod-severe TBI | No RHI exposure |
|---|---|---|---|---|---|---|---|---|---|
| **Delayed onset (of one or more core clinical features)** | 11 (44) | 28 (75.7) | 5 (29.4) | 16 (76.2) | 3 (15.8) | 9 (60) | 3 (20) | 0 | 0 |
| **Parkinsonism** | 6 (24) | 14 (37.8) | 2 (11.8) | 7 (33.3) | 5 (26.3) | 2 (13.3) | 3 (20) | 2 (13.3) | 7 (23.3) |
| **Other Motor Signs** | 0 | 2 (5.4) | 0 | 4 (19) | 0 | 1 (6.7) | 0 | 3 (20) | 1 (3.3) |
| **Motor Neuron Disease** | 2 (8) | 1 (2.7) | 1 (5.9) | 1 (4.8) | 4 (21.1) | 3 (20) | 4 (26.7) | 0 | 1 (3.3) |
| **Psychiatric Features** | 18 (72) | 36 (97.3) | 16 (94.1) | 19 (90.5) | 13 (68.4) | 15 (100) | 12 (80) | 11 (73.3) | 21 (70) |
| Anxiety | 11 (44) | 24 (64.9) | 8 (47.1) | 13 (61.9) | 5 (26.3) | 12 (80) | 7 (46.7) | 3 (20) | 9 (30) |
| Apathy | 16 (64) | 29 (78.4) | 10 (58.8) | 17 (81) | 11 (57.9) | 9 (60) | 5 (33.3) | 8 (53.3) | 17 (56.7) |
| Depression | 14 (56) | 25 (67.6) | 14 (82.4) | 13 (61.9) | 10 (52.6) | 13 (86.7) | 10 (66.7) | 6 (40) | 13 (43.3) |
| Paranoia | 11 (44) | 20 (54.1) | 7 (41.2) | 12 (57.1) | 5 (26.3) | 7 (46.7) | 4 (26.7) | 5 (33.3) | 5 (16.7) |
| **Additional clinical information** | | | | | | | | | |
| Neuropsychological testing | 14 (56) | 29 (78.4) | 8 (47.1) | 11 (52.4) | 13 (68.4) | 4 (26.7) | 10 (66.7) | 11 (73.3) | 23 (79.3) |
| MRI | 5 (20) | 2 (5.4) | 5 (29.4) | 7 (33.3) | 6 (31.6) | 3 (20) | 6 (40) | 6 (40) | 12 (41.4) |
| AD biomarkers (either CSF or amyloid PET) | 0 | 3 (8.1) | 0 | 2 (9.5) | 1 (5.3) | 0 | 0 | 0 | 2 (6.9) |
| **Neuropathology** | | | | | | | | | |
| **CTE** | 4 (16) | 29 (78.4) | 4 (23.5) | 11 (52.4) | 1 (5.3) | 4 (26.7) | 2 (13.3) | 2 (13.3) | 0 |
| **CTE Stage** | | | | | | | | | |
| Stage I | 2 (8) | 2 (5.4) | 3 (17.6) | 2 (9.5) | 1 (5.3) | 1 (6.7) | 1 (6.7) | 2 (13.3) | 0 |
| Stage II | 1 (4) | 6 (5.4) | 1 (5.9) | 1 (4.8) | 0 | 2 (13.3) | 0 | 0 | 0 |
| Stage III | 1 (4) | 10 (27) | 0 | 3 (14.3) | 0 | 1 (6.7) | 1 (6.7) | 0 | 0 |
| Stage IV | 0 | 11 (29.7) | 0 | 5 (23.8) | 0 | 0 | 0 | 0 | 0 |
| **CTE II-IV** | 2 (8) | 27 (73) | 1 (5.9) | 9 (42.9) | 0 | 3 (20) | 1 (6.7) | 0 | 0 |
| **Alzheimer's disease NIA-Reagan criteria** | 5 (20) | 14 (37.8) | 5 (29.4) | 6 (28.6) | 7 (36.8) | 5 (33.3) | 7 (46.7) | 4 (26.7) | 16 (55.2) |
| **FTLD diagnosis** | 1 (4) | 3 (8.1) | 3 (17.6) | 1 (4.8) | 2 (10.5) | 1 (6.7) | 2 (13.3) | 1 (6.7) | 3 (10.3) |

| | College FB< | College FB+ | College sport< | College Sport+ | Military w/combat, no sport | Military w/combat+ sport | 1+mTBI w/LOC | 1+ mod-severe TBI | No RHI exposure |
|---|---|---|---|---|---|---|---|---|---|
| **Lewy body disease** | | | | | | | | | |
| Brainstem predominant | 2 (8) | 2 (5.4) | 2 (11.8) | 1 (4.8) | 2 (10.5) | 0 | 1 (6.7) | 2 (13.3) | 5 (17.2) |
| Limbic (transitional) | 0 | 3 (8.1) | 0 | 2 (9.5) | 1 (5.3) | 1 (6.7) | 2 (13.3) | 1 (6.7) | 1 (3.4) |
| Neocortical (diffuse) | 0 | 0 | 0 | 0 | 1 (5.3) | 0 | 0 | 1 (6.7) | 1 (3.4) |
| Olfactory bulb | 1 (4) | 0 | 0 | 0 | 0 | 0 | 0 | 0 | 0 |
| **Motor neuron disease** | | | | | | | | | |
| With TDP-43 inclusions in motor neurons | 2 (8) | 1 (2.7) | 1 (5.9) | 1 (4.8) | 4 (21.1) | 3 (20) | 3 (20) | 0 | 1 (3.4) |
| With other inclusions | 0 | 0 | 0 | 0 | 0 | 0 | 1 (6.7) | 0 | 0 |
| **Thal phase (n=163)** | | | | | | | | | |
| Phase 1 (A1) | 2 (9.5) | 2 (5.6) | 1 (8.3) | 0 | 2 (18.2) | 1 (6.7) | 4 (28.6) | 1 (9.1) | 2 (8.7) |
| Phase 2 (A1) | 1 (4.8) | 1 (2.8) | 0 | 0 | 0 | 0 | 0 | 0 | 3 (13) |
| Phase 3 (A2) | 2 (9.5) | 5 (13.9) | 0 | 3 (15) | 3 (27.3) | 2 (13.3) | 4 (28.6) | 2 (18.2) | 2 (8.7) |
| Phase 4 (A3) | 2 (9.5) | 6 (16.7) | 0 | 5 (25) | 3 (27.3) | 0 | 2 (14.3) | 4 (36.4) | 6 (26.1) |
| Phase 5 (A3) | 1 (4.8) | 6 (16.7) | 1 (8.3) | 3 (15) | 0 | 3 (20) | 2 (14.3) | 0 | 6 (26.1) |
| **Braak stage (n=180)** | | | | | | | | | |
| Stage I (B1) | 8 (32) | 4 (11.1) | 1 (6.7) | 3 (14.3) | 0 | 1 (6.7) | 1 (6.7) | 3 (20) | 1 (4.2) |
| Stage II (B1) | 4 (16) | 1 (2.8) | 0 | 0 | 2 (14.3) | 2 (13.3) | 3 (20) | 1 (6.7) | 3 (12.5) |
| Stage III (B2) | 1 (4) | 3 (8.3) | 1 (6.7) | 4 (19) | 4 (28.6) | 0 | 4 (26.7) | 3 (20) | 4 (16.7) |
| Stage IV (B2) | 2 (8) | 5 (13.9) | 1 (6.7) | 1 (4.8) | 1 (7.1) | 2 (13.3) | 2 (13.3) | 2 (13.3) | 1 (4.2) |
| Stage V (B3) | 2 (8) | 9 (25) | 2 (13.3) | 2 (9.5) | 0 | 0 | 1 (6.7) | 1 (6.7) | 9 (37.5) |
| Stage VI (B3) | 1 (4) | 3 (8.3) | 1 (6.7) | 4 (19) | 1 (7.1) | 3 (20) | 2 (13.3) | 3 (20) | 5 (20.8) |
| **CERAD score (neuritic plaques) (n=180)** | | | | | | | | | |
| Sparse neuritic plaques (C1) | 5 (20) | 6 (16.7) | 2 (13.3) | 3 (14.3) | 3 (21.4) | 1 (6.7) | 5 (33.3) | 4 (26.7) | 4 (16.7) |
| Moderate neuritic plaques (C2) | 3 (12) | 7 (19.4) | 0 | 3 (14.3) | 2 (14.3) | 1 (6.7) | 4 (26.7) | 3 (20) | 7 (29.2) |
| Frequent neuritic plaques (C3) | 1 (4) | 2 (5.6) | 1 (6.7) | 3 (14.3) | 1 (7.1) | 3 (20) | 0 | 1 (6.7) | 7 (29.2) |

| | College FB< | College FB+ | College sport< | College Sport+ | Military w/combat, no sport | Military w/combat+ sport | 1+mTBI w/LOC | 1+ mod-severe TBI | No RHI exposure |
|---|---|---|---|---|---|---|---|---|---|
| **CERAD score (diffuse plaques) (n=180)**<br>Sparse diffuse plaques<br>Moderate diffuse plaques<br>Frequent diffuse plaques | 5 (20)<br>2 (8)<br>4 (16) | 4 (11.1)<br>8 (22.2)<br>8 (22.2) | 2 (13.3)<br>0<br>3 (20) | 1 (4.8)<br>5 (23.8)<br>6 (28.6) | 2 (14.3)<br>4 (28.6)<br>4 (28.6) | 1 (6.7)<br>1 (6.7)<br>4 (26.7) | 5 (33.3)<br>5 (33.3)<br>3 (20) | 3 (20)<br>2 (13.3)<br>3 (20) | 3 (12.5)<br>7 (29.2)<br>10 (41.7) |
| **Arteriolosclerosis (n=180)**<br>Mild<br>Moderate<br>Severe | 9 (36)<br>6 (24)<br>4 (16) | 14 (38.9)<br>13 (36.1)<br>5 (13.9) | 5 (33.3)<br>3 (20)<br>2 (13.3) | 5 (23.8)<br>7 (33.3)<br>3 (14.3) | 4 (28.6)<br>4 (28.6)<br>4 (28.6) | 6 (40)<br>4 (26.7)<br>0 | 3 (20)<br>9 (60)<br>2 (13.3) | 6 (40)<br>4 (26.7)<br>2 (13.3) | 12 (50)<br>8 (33.3)<br>3 (12.5) |
| **Atherosclerosis (n=177)**<br>Mild<br>Moderate<br>Severe | 7 (28)<br>4 (16)<br>0 | 9 (25.7)<br>6 (17.1)<br>1 (2.9) | 5 (33.3)<br>0<br>0 | 2 (10.5)<br>3 (15.8)<br>0 | 6 (42.9)<br>1 (7.1)<br>2 (14.3) | 3 (20)<br>1 (6.7)<br>1 (6.7) | 8 (53.3)<br>1 (6.7)<br>2 (13.3) | 6 (40)<br>3 (20)<br>1 (6.7) | 8 (33.3)<br>2 (8.3)<br>4 (16.7) |
| **White Matter Rarefaction (n=146)**<br>Mild<br>Moderate<br>Severe | 12 (54.5)<br>3 (13.6)<br>0 | 12 (34.3)<br>15 (42.9)<br>2 (5.7) | 4 (30.8)<br>2 (15.4)<br>1 (7.7) | 6 (30)<br>10 (50)<br>1 (5) | 5 (38.5)<br>3 (23.1)<br>1 (7.7) | 9 (69.2)<br>2 (15.4)<br>0 | 5 (38.5)<br>7 (53.8)<br>1 (6.7) | 1 (6.7)<br>2 (13.3)<br>0 | 6 (54.5)<br>1 (9.1)<br>3 (27.3) |
| **Cerebral Amyloid Angiopathy (CAA) (n=180)**<br>Mild<br>Moderate<br>Severe | 5 (20)<br>0<br>3 (12) | 4 (11.1)<br>3 (8.3)<br>3 (8.3) | 2 (13.3)<br>1 (6.7)<br>1 (6.7) | 2 (9.5)<br>4 (19)<br>0 | 2 (14.3)<br>6 (42.9)<br>1 (7.1) | 2 (13.3)<br>4 (26.7)<br>0 | 4 (26.7)<br>2 (13.3)<br>1 (6.7) | 3 (20)<br>2 (13.3)<br>1 (6.7) | 4 (16.7)<br>4 (16.7)<br>6 (25) |
| **Old microinfarcts (n=180)** | 6 (24) | 6 (16.7) | 4 (26.7) | 2 (9.5) | 3 (21.4) | 3 (20) | 6 (40) | 5 (33.3) | 4 (16.7) |
| **Old cerebral microbleeds (n=163)** | 2 (9.5) | 0 | 1 (8.3) | 1 (5) | 0 | 0 | 1 (7.1) | 4 (36.4) | 3 (13) |

| | College FB< | College FB+ | College sport< | College Sport+ | Military w/combat, no sport | Military w/combat+ sport | 1+mTBI w/LOC | 1+ mod-severe TBI | No RHI exposure |
|---|---|---|---|---|---|---|---|---|---|
| **TDP-43 inclusions (n=181)** | 2 (8) | 12 (33.3) | 2 (13.3) | 4 (19) | 4 (26.7) | 5 (33.3) | 6 (40) | 3 (20) | 9 (37.5) |
| Amygdala | 0 | 11 (29.7) | 2 (11.8) | 3 (14.3) | 2 (10.5) | 4 (26.7) | 3 (20) | 1 (6.7) | 8 (27.6) |
| Hippocampus | 0 | 11 (29.7) | 2 (11.8) | 3 (14.3) | 2 (10.5) | 5 (33.3) | 3 (20) | 2 (13.3) | 5 (17.2) |
| Neocortex | 2 (8) | 7 (18.9) | 1 (5.9) | 3 (14.3) | 3 (15.8) | 3 (20) | 2 (13.3) | 3 (20) | 0 |

**eTable 4. Descriptives and Neuropathology (stratified by age bins)**

| | 20-34 | 35-49 | 50-64 | 65-79 | 80+ |
|---|---|---|---|---|---|
| **Number** | 25 | 25 | 29 | 43 | 71 |
| **Demographics** | | | | | |
| Age at death (years) | 27.96 ± 4.15 | 42.36 ± 3.76 | 58.31 ± 4.29 | 72.4 ± 4.41 | 87.9 ± 6.29 |
| Female sex | 0 | 0 | 4 (13.8) | 7 (16.3) | 29 (40.8) |
| Race | | | | | |
| Black | 4 (16) | 2 (8) | 5 (17.2) | 1 (2.3) | 3 (4.2) |
| White | 16 (64) | 21 (84) | 24 (82.8) | 35 (81.4) | 62 (87.3) |
| Multiple | 4 (16) | 1 (4) | 0 | 0 | 0 |
| Other | 0 | 1 (4) | 0 | 0 | 0 |
| Unreported | 1 (4) | 0 | 0 | 7 (16.3) | 6 (8.4) |
| **Donor Recruitment categories** | | | | | |
| RHI exposure | | | | | |
| College football< | 7 (28) | 3 (12) | 4 (13.8) | 6 (14) | 5 (7) |
| College football+ | 8 (32) | 3 (12) | 8 (27.6) | 8 (18.6) | 10 (14.1) |
| College contact sport< | 4 (16) | 6 (24) | 0 | 2 (4.7) | 5 (7) |
| College contact sport+ | 2 (8) | 5 (20) | 8 (27.6) | 1 (2.3) | 5 (7) |
| Military w/combat, no sport | 0 | 2 (8) | 1 (3.4) | 5 (11.6) | 11 (15.5) |
| Military w/combat + sport | 2 (8) | 6 (24) | 2 (6.9) | 4 (9.3) | 1 (1.4) |
| 1+ mTBI w/LOC, no sport/military | 0 | 0 | 3 (10.3) | 3 (7) | 9 (12.7) |
| 1+ mod-severe TBI, no sport/military | 2 (8) | 0 | 3 (10.3) | 3 (7) | 7 (9.9) |
| No RHI exposure | 0 | 0 | 0 | 11 (25.6) | 18 (25.4) |
| Brain Bank | | | | | |
| UNITE | 24 (96) | 22 (88) | 16 (55.2) | 11 (25.6) | 13 (18.3) |
| FHS | 0 | 0 | 5 (17.2) | 3 (7) | 24 (33.8) |
| BU ADRC | 0 | 0 | 1 (3.4) | 6 (14) | 18 (25.4) |
| Mt. Sinai ADRC | 0 | 0 | 0 | 7 (16.3) | 9 (12.7) |
| VA ALS | 0 | 0 | 4 (13.8) | 8 (18.6) | 4 (5.6) |
| LETBI | 1 (4) | 3 (12) | 3 (10.3) | 8 (18.6) | 3 (4.2) |

| | 20-34 | 35-49 | 50-64 | 65-79 | 80+ |
|---|---|---|---|---|---|
| **TES Criteria** | | | | | |
| **TES status** | | | | | |
| No | 11 (44) | 10 (40) | 14 (48.3) | 31 (72.1) | 55 (77.5) |
| Suggestive | 7 (28) | 4 (16) | 0 | 2 (4.7) | 2 (2.8) |
| Possible | 6 (24) | 8 (32) | 6 (20.7) | 4 (9.3) | 4 (5.6) |
| Probable | 1 (4) | 3 (12) | 9 (31) | 6 (14) | 10 (14.1) |
| **Substantial exposure to RHI** | 24 (96) | 24 (96) | 27 (93.1) | 23 (53.5) | 31 (43.7) |
| **Extensive exposure to RHI** | 7 (28) | 5 (20) | 9 (31) | 6 (14) | 10 (14.1) |
| **Cognitive impairment** | 10 (40) | 20 (80) | 22 (75.9) | 33 (76.7) | 58 (81.7) |
| **Neurobehavioral dysregulation** | 24 (96) | 23 (92) | 20 (69) | 24 (55.8) | 25 (35.2) |
| **Progressive course** | | | | | |
| Yes | 19 (76) | 23 (92) | 23 (79.3) | 35 (81.4) | 58 (81.7) |
| No | 1 (4) | 0 | 0 | 0 | 0 |
| N/A | 1 (4) | 0 | 6 (20.7) | 7 (16.3) | 12 (16.9) |
| Inconclusive | 4 (16) | 2 (8) | 0 | 1 (2.3) | 1 (1.4) |
| **Fully Accounted for by Other Disorders** | | | | | |
| No | 17 (68) | 17 (68) | 15 (51.7) | 12 (27.9) | 19 (26.8) |
| Yes | 7 (28) | 7 (28) | 7 (24.1) | 20 (46.5) | 31 (43.7) |
| N/A | 1 (4) | 1 (4) | 7 (24.1) | 11 (25.6) | 21 (29.6) |
| **Level of Functional Dependence/Dementia** | | | | | |
| Independent/no dementia | 10 (40) | 3 (12) | 7 (24.1) | 7 (16.3) | 13 (18.3) |
| Subtle functional limitation | 15 (60) | 19 (76) | 4 (13.8) | 3 (7) | 6 (8.5) |
| Mild dementia | 0 | 3 (12) | 7 (24.1) | 4 (9.3) | 8 (11.3) |
| Moderate dementia | 0 | 0 | 5 (17.2) | 5 (11.6) | 13 (18.3) |
| Severe dementia | 0 | 0 | 6 (20.7) | 24 (55.8) | 31 (43.7) |
| **Delayed onset (of one or more core clinical features)** | 7 (28) | 13 (52) | 18 (62.1) | 17 (39.5) | 20 (28.2) |
| **Parkinsonism** | 1 (4) | 1 (4) | 9 (31) | 17 (19.5) | 20 (28.2) |
| **Other Motor Signs** | 0 | 0 | 2 (6.9) | 2 (4.7) | 7 (9.9) |
| **Motor Neuron Disease** | 0 | 0 | 5 (17.2) | 8 (18.6) | 4 (5.6) |

| | 20-34 | 35-49 | 50-64 | 65-79 | 80+ |
|---|---|---|---|---|---|
| **Psychiatric Features** | 24 (96) | 25 (100) | 23 (79.3) | 36 (83.7) | 53 (74.6) |
| Anxiety | 17 (68) | 21 (84) | 18 (62.1) | 18 (41.9) | 18 (25.4) |
| Apathy | 20 (80) | 18 (72) | 18 (62.1) | 29 (67.4) | 37 (52.1) |
| Depression | 22 (88) | 24 (96) | 19 (65.5) | 20 (46.5) | 33 (46.5) |
| Paranoia | 17 (68) | 15 (60) | 12 (41.4) | 15 (34.9) | 17 (23.9) |
| **Additional clinical information** | | | | | |
| Neuropsychological testing | 3 (12) | 11 (44) | 19 (65.5) | 26 (60.5) | 64 (90.1) |
| MRI | 2 (8) | 4 (16) | 11 (37.9) | 14 (32.6) | 21 (29.6) |
| AD biomarkers (either CSF or amyloid PET) | 0 | 1 (4) | 2 (6.9) | 2 (4.7) | 3 (4.2) |
| **Neuropathology** | | | | | |
| **CTE** | 8 (32) | 12 (48) | 16 (55.2) | 7 (16.3) | 14 (19.7) |
| **CTE Stage** | | | | | |
| Stage I | 2 (8) | 6 (24) | 4 (13.8) | 0 | 2 (2.8) |
| Stage II | 5 (20) | 4 (16) | 1 (3.4) | 0 | 1 (1.4) |
| Stage III | 1 (4) | 1 (4) | 8 (27.6) | 4 (9.3) | 2 (2.8) |
| Stage IV | 0 | 1 (4) | 3 (10.3) | 3 (7) | 9 (12.7) |
| **CTE II-IV** | 6 (24) | 6 (24) | 12 (41.4) | 7 (16.3) | 12 (16.9) |
| **Alzheimer's disease NIA-Reagan criteria** | 0 | 0 | 8 (27.6) | 24 (55.8) | 37 (52.1) |
| **FTLD diagnosis** | 0 | 0 | 1 (3.4) | 5 (11.6) | 11 (15.5) |
| **Lewy body disease** | | | | | |
| Brainstem predominant | 0 | 1 (4) | 2 (6.9) | 5 (11.6) | 9 (12.7) |
| Limbic (transitional) | 0 | 0 | 2 (6.9) | 4 (9.3) | 5 (7) |
| Neocortical (diffuse) | 0 | 0 | 0 | 1 (2.3) | 2 (2.8) |
| Olfactory bulb | 0 | 0 | 0 | 1 (2.3) | 0 |
| **Motor neuron disease** | | | | | |
| With TDP-43 inclusions in motor neurons | 0 | 0 | 5 (17.2) | 7 (16.3) | 4 (5.6) |
| With other inclusions | 0 | 0 | 0 | 1 (2.3) | 0 |
| **Thal phase (n=163)** | | | | | |
| Phase 1 (A1) | 0 | 1 (4) | 1 (4) | 6 (15.4) | 7 (14.3) |
| Phase 2 (A1) | 0 | 0 | 0 | 1 (2.6) | 4 (8.2) |
| Phase 3 (A2) | 0 | 1 (4) | 3 (12) | 9 (23.1) | 10 (20.4) |
| Phase 4 (A3) | 0 | 0 | 3 (12) | 8 (20.5) | 17 (34.7) |
| Phase 5 (A3) | 0 | 0 | 5 (20) | 11 (28.2) | 6 (12.2) |

| | 20-34 | 35-49 | 50-64 | 65-79 | 80+ |
|---|---|---|---|---|---|
| **Braak stage (n=180)** | | | | | |
| Stage I (B1) | 2 (8) | 5 (20) | 8 (27.6) | 4 (10) | 3 (4.9) |
| Stage II (B1) | 0 | 0 | 4 (13.8) | 4 (10) | 8 (13.1) |
| Stage III (B2) | 0 | 1 (4) | 4 (13.8) | 2 (5) | 17 (27.9) |
| Stage IV (B2) | 0 | 1 (4) | 1 (3.4) | 6 (15) | 9 (14.8) |
| Stage V (B3) | 0 | 0 | 3 (10.3) | 13 (32.5) | 10 (16.4) |
| Stage VI (B3) | 0 | 0 | 6 (20.7) | 5 (12.5) | 12 (19.7) |
| **CERAD score (neuritic plaques) (n=180)** | | | | | |
| Sparse neuritic plaques (C1) | 0 | 0 | 3 (10.3) | 12 (30) | 18 (29.5) |
| Moderate neuritic plaques (C2) | 0 | 0 | 4 (13.8) | 7 (17.5) | 19 (31.1) |
| Frequent neuritic plaques (C3) | 0 | 0 | 3 (10.3) | 11 (27.5) | 5 (8.2) |
| **CERAD score (diffuse plaques) (n=180)** | | | | | |
| Sparse diffuse plaques | 0 | 1 (4) | 1 (3.4) | 11 (27.5) | 13 (21.3) |
| Moderate diffuse plaques | 0 | 1 (4) | 6 (20.7) | 10 (25) | 17 (27.9) |
| Frequent diffuse plaques | 0 | 0 | 6 (20.7) | 14 (35) | 25 (41) |
| **Arteriolosclerosis (n=180)** | | | | | |
| Mild | 8 (32) | 9 (36) | 10 (34.5) | 18 (45) | 19 (31.1) |
| Moderate | 4 (16) | 4 (16) | 9 (31) | 15 (37.5) | 26 (42.6) |
| Severe | 0 | 2 (8) | 2 (6.9) | 6 (15) | 15 (24.6) |
| **Atherosclerosis (n=177)** | | | | | |
| Mild | 1 (4.3) | 1 (4.2) | 9 (31) | 19 (47.5) | 24 (39.3) |
| Moderate | 0 | 0 | 2 (6.9) | 4 (10) | 15 (24.6) |
| Severe | 0 | 0 | 0 | 3 (7.5) | 8 (13.1) |
| **White Matter Rarefaction (n=146)** | | | | | |
| Mild | 10 (41.7) | 11 (50) | 10 (38.5) | 10 (40) | 19 (38.8) |
| Moderate | 5 (20.8) | 5 (22.7) | 9 (34.6) | 8 (32) | 18 (36.7) |
| Severe | 0 | 0 | 0 | 2 (8) | 7 (14.3) |
| **Cerebral Amyloid Angiopathy (CAA) (n=180)** | | | | | |
| Mild | 1 (4) | 0 | 4 (13.8) | 9 (22.5) | 14 (23) |
| Moderate | 0 | 0 | 3 (10.3) | 6 (15) | 17 (27.9) |
| Severe | 0 | 0 | 2 (6.9) | 7 (17.5) | 7 (11.5) |
| **Old microinfarcts (n=180)** | 1 (4) | 2 (8) | 2 (6.9) | 12 (30) | 22 (36.1) |
| **Old cerebral microbleeds (n=163)** | 1 (4) | 2 (8) | 3 (12) | 2 (5.1) | 4 (8.2) |

| | 20-34 | 35-49 | 50-64 | 65-79 | 80+ |
|---|---|---|---|---|---|
| **TDP-43 inclusions (n=181)** | 0 | 0 | 9 (31) | 18 (45) | 20 (32.3) |
| Amygdala | 0 | 0 | 6 (20.7) | 15 (34.9) | 13 (18.3) |
| Hippocampus | 0 | 0 | 7 (24.1) | 13 (30.2) | 13 (18.3) |
| Neocortex | 0 | 0 | 7 (24.1) | 10 (23.3) | 7 (9.9) |

### eTable 5. TES consensus diagnosis by CTE pathology sensitivity analyses

**CTE stage II-IV vs. $CTE_{pos/prob}$ (presence of substance use disorder (n=56))**

| | CTE stage II-IV | No CTE/CTE stage I | Totals |
|---|---|---|---|
| $CTE_{pos/prob}$ | 8 | 11 | 19 |
| no TES/$CTE_{sug}$ | 6 | 31 | 37 |
| Totals | 14 | 42 | 56 |
| | Sensitivity: 0.57 (0.31,0.83) | Specificity: 0.74 (0.61,0.87) | |

Positive likelihood ratio: 2.18 (0.98, 4.88)
Negative likelihood ratio: 0.58 (0.27, 1.24)

**CTE stage II-IV vs. $CTE_{pos/prob}$ (absence of substance use disorder (n=137))**

| | CTE stage II-IV | No CTE/CTE stage I | Totals |
|---|---|---|---|
| $CTE_{pos/prob}$ | 25 | 13 | 38 |
| no TES/$CTE_{sug}$ | 4 | 95 | 99 |
| Totals | 29 | 108 | 137 |
| | Sensitivity: 0.86 (0.74,0.99) | Specificity: 0.88 (0.82,0.94) | |

Positive likelihood ratio: 7.16 (3.96, 12.97)
Negative likelihood ratio: 0.16 (0.06, 0.41)

**CTE stage II-IV vs. $CTE_{pos/prob}$ (presence of mental health disorder(s) (n=88))**

| | CTE stage II-IV | No CTE/CTE stage I | Totals |
|---|---|---|---|
| $CTE_{pos/prob}$ | 12 | 15 | 27 |
| no TES/$CTE_{sug}$ | 7 | 54 | 61 |
| Totals | 19 | 69 | 88 |
| | Sensitivity: 0.63 (0.42,0.85) | Specificity: 0.78 (0.69,0.88) | |

Positive likelihood ratio: 2.91 (1.51, 5.60)
Negative likelihood ratio: 0.47 (0.24, 0.94)

**CTE stage II-IV vs. $CTE_{pos/prob}$ (absence of mental health disorder (n=105))**

| | CTE stage II-IV | No CTE/CTE stage I | Totals |
|---|---|---|---|
| $CTE_{pos/prob}$ | 21 | 9 | 30 |
| no TES/$CTE_{sug}$ | 3 | 72 | 875 |
| Totals | 24 | 89 | 105 |
| | Sensitivity: 0.88 (0.74,1) | Specificity: 0.89 (0.82,0.96) | |

Positive likelihood ratio: 7.88 (3.89, 15.94)
Negative likelihood ratio: 0.14 (0.05, 0.42)

**CTE stage II-IV vs. $CTE_{pos/prob}$ (Neuropsychological testing data available (n=123))**

| | CTE stage II-IV | No CTE/CTE stage I | Totals |
|---|---|---|---|
| $CTE_{pos/prob}$ | 27 | 11 | 38 |
| no TES/$CTE_{sug}$ | 3 | 82 | 85 |
| Totals | 30 | 93 | 123 |
| | Sensitivity: 0.9 (0.79,1) | Specificity: 0.88 (0.82,0.95) | |

Positive likelihood ratio: 7.61 (4.03, 14.38)
Negative likelihood ratio: 0.11 (0.04, 0.35)

**CTE stage II-IV vs. $CTE_{pos/prob}$ (participants with no football exposure (n=122))**

| | CTE stage II-IV | No CTE/CTE stage I | Totals |
|---|---|---|---|
| $CTE_{pos/prob}$ | 10 | 10 | 20 |
| no TES/$CTE_{sug}$ | 2 | 100 | 102 |
| Totals | 12 | 110 | 122 |
| | Sensitivity: 0.83 (0.62,1) | Specificity: 0.91 (0.86,0.96) | |

Positive likelihood ratio: 9.17 (4.57, 18.37)
Negative likelihood ratio: 0.18 (0.05, 0.69)

**CTE stage II-IV vs. $CTE_{pos/prob}$ (neuropathological presence of Alzheimer's disease (n=69))**

| | CTE stage II-IV | No CTE/CTE stage I | Totals |
|---|---|---|---|
| $CTE_{pos/prob}$ | 14 | 9 | 23 |
| no TES/$CTE_{sug}$ | 2 | 44 | 46 |
| Totals | 16 | 53 | 69 |
| | Sensitivity: 0.88 (0.71,1) | Specificity: 0.83 (0.73,0.93) | |

Positive likelihood ratio: 5.15 (2.48, 10.70)
Negative likelihood ratio: 0.15 (0.04 ,0.58)

**CTE stage II-IV vs. $CTE_{pos/prob}$ (diagnosis of another non-CTE and non-AD neurodegenerative pathology (n=57))**

| | CTE stage II-IV | No CTE/CTE stage I | Totals |
|---|---|---|---|
| $CTE_{pos/prob}$ | 13 | 5 | 18 |
| no TES/$CTE_{sug}$ | 0 | 39 | 39 |
| Totals | 13 | 44 | 57 |
| | Sensitivity: 1 | Specificity: 0.89 (0.79,0.98) | |

Positive likelihood ratio: 8.8 (3.49,22.19)
Negative likelihood ratio: 0

**CTE stage II-IV vs. Substantial or Extensive RHI Exposure**

| | CTE stage II-IV | NoCTE/CTE I | Totals |
|---|---|---|---|
| Substantial/ Extensive RHI | 42 | 87 | 129 |
| Not Substantial/ Extensive RHI | 1 | 63 | 64 |
| Totals | 43 | 150 | 193 |
| | Sensitivity: 0.98 (0.93,1) | Specificity: 0.42 (0.34,0.50) | |

Positive likelihood ratio: 1.68 (1.29, 2.20)
Negative likelihood ratio: 0.06 (0.01, 0.39)

**CTE stage II-IV vs. Substantial or Extensive RHI Exposure, plus cognitive impairment**

| | CTE stage II-IV | NoCTE/CTE I | Totals |
|---|---|---|---|
| Substantial/ Extensive RHI + cognitive impairment | 35 | 57 | 92 |
| Not Substantial/ Extensive RHI + cognitive impairment | 8 | 93 | 101 |
| Totals | 43 | 150 | 193 |
| | Sensitivity: 0.81 (0.70,0.93) | Specificity: 0.62 (0.54,0.70) | |

Positive likelihood ratio: 2.14 (1.53, 3)
Negative likelihood ratio: 0.30 (0.15, 0.59)

**CTE stage II-IV vs. Extensive RHI Exposure**

| | CTE stage II-IV | NoCTE/CTEI | Totals |
|---|---|---|---|
| Extensive RHI | 30 | 6 | 36 |
| Not Extensive RHI | 13 | 144 | 157 |
| Totals | 43 | 150 | 193 |
| | Sensitivity: 0.70 (0.54,0.83) | Specificity: 0.96 (0.92,0.99) | |

Positive likelihood ratio: 17.4 (7.77,39.14)
Negative likelihood ratio: 0.31 (0.20,0.50)

**eTable 6. TES criteria, stratified by CTE Stage**

| | No CTE | CTE I | CTE II | CTEIII | CTE IV |
|---|---|---|---|---|---|
| **Number (%)** | 136 | 14 | 11 | 16 | 16 |
| **Age at death (years), SD** | 69.47 ± 22.39 | 49.93 ± 17.57 | 42.91 ± 17.77 | 62.13 ± 13.73 | 74.38 ± 12.53 |
| **TES status** | | | | | |
| No | 106 (77.9) | 8 (57.1) | 5 (45.5) | 2 (12.5) | 0 |
| Suggestive | 11 (8.1) | 1 (7.1) | 3 (27.3) | 0 | 0 |
| Possible | 16 (11.8) | 4 (28.6) | 1 (9.1) | 5 (31.3) | 2 (12.5) |
| Probable | 3 (2.2) | 1 (7.1) | 2 (18.2) | 9 (56.3) | 14 (87.5) |
| **Cognitive impairment** | 96 (70.6) | 11 (78.6) | 5 (45.5) | 15 (93.8) | 16 (100) |
| **Neurobehavioral dysregulation** | 67 (49.3) | 12 (85.7) | 9 (81.8) | 14 (87.5) | 14 (87.5) |
| **Progressive course** | | | | | |
| Yes | 106 (77.9) | 12 (85.7) | 9 (81.8) | 15 (93.8) | 16 (100) |
| No | 1 (0.7) | 0 | 0 | 0 | 0 |
| N/A | 23 (16.9) | 1 (7.1) | 1 (9.1) | 1 (6.3) | 0 |
| Inconclusive | 6 (4.4) | 1 (7.1) | 1 (9.1) | 0 | 0 |
| **Fully Accounted for by Other Disorders** | | | | | |
| No | 36 (26.5) | 7 (50) | 7 (63.6) | 14 (87.5) | 16 (100) |
| Yes | 63 (46.3) | 4 (42.9) | 2 (18.2) | 1 (6.3) | 0 |
| N/A | 37 (27.2) | 1 (7.1) | 2 (18.2) | 1 (6.3) | 0 |
| **Level of Functional Dependence/Dementia** | | | | | |
| Independent/no dementia | 33 (24.3) | 3 (21.4) | 3 (27.3) | 1 (6.3) | 0 |
| Subtle functional limitation | 28 (20.6) | 7 (50) | 7 (63.6) | 4 (25) | 1 (6.3) |
| Mild dementia | 15 (11) | 1 (7.1) | 1 (9.1) | 4 (25) | 1 (6.3) |
| Moderate dementia | 14 (10.3) | 2 (14.3) | 0 | 2 (12.5) | 5 (31.3) |
| Severe dementia | 46 (33.8) | 1 (7.1) | 0 | 5 (31.3) | 9 (56.3) |
| **Delayed onset (of one or more core clinical features)** | 34 (25) | 6 (42.9) | 6 (54.5) | 13 (81.3) | 16 (100) |
| **Parkinsonism** | 28 (20.6) | 4 (28.6) | 1 (9.1) | 6 (37.5) | 9 (56.3) |
| **Other Motor Signs** | 6 (4.4) | 1 (7.1) | 0 | 1 (6.3) | 3 (18.8) |
| **Motor Neuron Disease** | 11 (8.1) | 3 (21.4) | 1 (9.1) | 1 (6.3) | 1 (6.3) |

| | | | | | |
|---|---|---|---|---|---|
| **Psychiatric Features** | 108 (79.4) | 11 (78.6) | 10 (90.9) | 16 (100) | 16 (100) |
| Anxiety | 57 (41.9) | 7 (50) | 7 (63.6) | 11 (68.8) | 10 (62.5) |
| Apathy | 79 (58.1) | 9 (64.3) | 7 (63.6) | 13 (81.3) | 14 (87.5) |
| Depression | 77 (56.6) | 10 (71.4) | 9 (81.8) | 13 (81.3) | 9 (56.3) |
| Paranoia | 45 (33.1) | 6 (42.9) | 5 (45.5) | 10 (62.5) | 10 (62.5) |

**eFigure 1. Standards for the Reporting of Diagnostic Accuracy Studies (STARD) flow diagram**

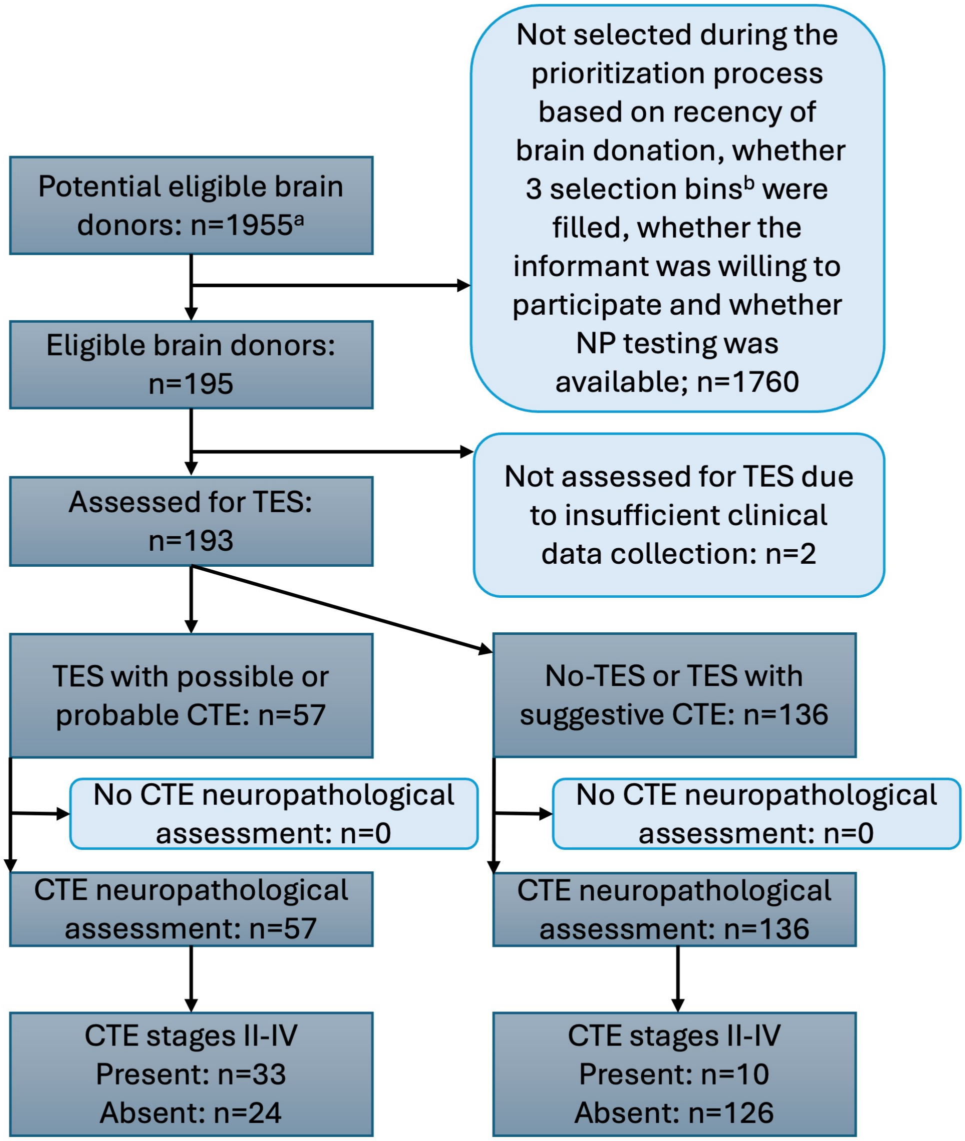


Legend: STARD flowchart showing study selection of brain donors and breakdown by results of index test and reference standard.

[a] Sample size for potential eligible brain donors is the total brain donors across all brain banks who had been donated prior to or during the study period and who had not been included in a previous study of TES.[10]

[b] Three selection bins are brain bank, age group and RHI/TBI exposure group. See methods for additional description of these bins.

Abbreviations: CTE: chronic traumatic encephalopathy; NP: neuropsychological; TES: traumatic encephalopathy syndrome.

### eFigure 2. Consensus checklist clinical diagnoses beyond TES

**Consensus Clinical Diagnosis**

Select all that apply.

Please select diagnoses based on whether the clinical syndrome fits diagnostic criteria, regardless of whether you think the underlying pathology or combination of pathologies are present.

- [ ] Mild Cognitive Impairment *(Check all that apply)*:
- [ ] Amnestic Multidomain Dementia *(Select one)*:
- [ ] Posterior Cortical Atrophy (PCA) (or primary visual presentation)
- [ ] Primary Progressive Aphasia (PPA) *(Select one)*:
- [ ] Behavioral Variant FTD (bvFTD)
- [ ] Parkinson's Disease/Lewy Body Disease *(Select one)*:
- [ ] Non-amnestic Multidomain Dementia, not PCA, PPA, DLB, or bvFTD, or vascular dementia
- [ ] Stroke/Vascular Impairment *(Check all that apply)*:

---

- [ ] Alcohol-related Dementia
- [ ] Rapidly Progressive Dementia
- [ ] Cognitively Impaired Non-MCI
- [ ] Major Depressive Disorder
- [ ] Bipolar Disorder *(Select one)*:
- [ ] Trauma- and Stressor-Related Disorders *(Check all that apply)*:
- [ ] Anxiety Disorder *(Check all that apply)*:
- [ ] Psychotic Disorder *(Select one)*:

- [ ] Intermittent Explosive Disorder
- [ ] ADHD
- [ ] Learning Disability
- [ ] Substance Use Disorder
- [ ] Other Psychiatric Illness:
- [ ] PCS
- [ ] Residual problems after moderate to severe TBI
- [ ] ALS/Motor Neuron Disease
- [ ] Progressive Supranuclear Palsy
- [ ] Corticobasal Syndrome
- [ ] Huntington's Disease
- [ ] Multiple Sclerosis
- [ ] Epilepsy
- [ ] Residual problems after CNS neoplasm
- [ ] OSA contributing to neurological and/or psychiatric impairment
- [ ] Chronic Pain contributing to neurological and/or psychiatric impairment
- [ ] Headache contributing to neurological and/or psychiatric impairment
- [ ] Other Medical Illness:
- [ ] Iatrogenic Impairment
- [ ] No neurological, psychiatric, or cognitive diagnosis

**Comments**

### References


1. Mez J, Solomon TM, Daneshvar DH, et al. Assessing clinicopathological correlation in chronic traumatic encephalopathy: rationale and methods for the UNITE study. *Alzheimers Res Ther*. 2015;7(1):62. doi:10.1186/s13195-015-0148-8

2. Au R, Seshadri S, Knox K, et al. The Framingham Brain Donation Program: neuropathology along the cognitive continuum. *Curr Alzheimer Res*. 2012;9(6):673-686. doi:10.2174/156720512801322609

3. Alosco ML, Morrison M, Au R, et al. Boston University Alzheimer's Disease Research Center Clinical Core: Infrastructure to facilitate research on post-traumatic Alzheimer's disease and related dementias. *Alzheimers Dement*. 2025;21(9):e70654. doi:10.1002/alz.70654

4. Fischer DL, Grinberg LT, Ahrendsen JT, et al. Celebrating neuropathology's contributions to Alzheimer's Disease Research Centers. *Alzheimers Dement*. 2025;21(10):e70734. doi:10.1002/alz.70734

5. Edlow BL, Keene CD, Perl DP, et al. Multimodal Characterization of the Late Effects of Traumatic Brain Injury: A Methodological Overview of the Late Effects of Traumatic Brain Injury Project. *J Neurotrauma*. 2018;35(14):1604-1619. doi:10.1089/neu.2017.5457

6. Brady CB, Trevor KT, Stein TD, et al. The Department of Veterans Affairs Biorepository Brain Bank: a national resource for amyotrophic lateral sclerosis research. *Amyotroph Lateral Scler Frontotemporal Degener*. 2013;14(7-8):591-597. doi:10.3109/21678421.2013.822516

7. Katz DI, Bernick C, Dodick DW, et al. National Institute of Neurological Disorders and Stroke Consensus Diagnostic Criteria for Traumatic Encephalopathy Syndrome. *Neurology*. 2021;96(18):848-863. doi:10.1212/WNL.0000000000011850

8. Mez J, Daneshvar DH, Abdolmohammadi B, et al. Duration of American Football Play and Chronic Traumatic Encephalopathy. *Ann Neurol*. 2020;87(1):116-131. doi:10.1002/ana.25611

9. Abdolmohammadi B, Tuz-Zahra F, Uretsky M, et al. Duration of Ice Hockey Play and Chronic Traumatic Encephalopathy. *JAMA Netw Open*. 2024;7(12):e2449106. doi:10.1001/jamanetworkopen.2024.49106

10. Mez J, Alosco ML, Daneshvar DH, et al. Validity of the 2014 traumatic encephalopathy syndrome criteria for CTE pathology. *Alzheimer's & Dementia*. 2021;n/a(n/a). doi:10.1002/alz.12338
